# SARS-CoV-2 and the Pandemic Surge in Invasive Group A Streptococcal Disease

**DOI:** 10.64898/2026.03.19.26348823

**Authors:** David N. Fisman, Clara Eunyoung Lee, Natalie J. Wilson, Michelle Barton-Forbes, Simran K. Mann, Ashleigh R. Tuite

## Abstract

**Background:** Multiple countries reported unprecedented increases in invasive group A streptococcal (iGAS) disease following widespread SARS-CoV-2 circulation. Whether this surge reflects reduced pathogen exposure during non-pharmaceutical interventions (“immunity debt”) or effects of SARS-CoV-2 infection on host immunity remains unresolved.

**Methods:** We conducted a population-based time-series analysis of weekly iGAS incidence in central Ontario, Canada (population ≈11 million) from March 2011 through March 2024 (676 weeks). Using negative binomial panel regression, we modeled acute (2-week lagged) and cumulative SARS-CoV-2 exposure while adjusting for seasonality, secular trends, age, and sex. Population attributable fractions (PAFs) were estimated by counterfactual prediction. Specificity was assessed through negative control analyses (influenza, RSV). The immunity debt hypothesis was evaluated using cumulative streptococcal exposure as a predictor of iGAS.

**Results:** Among 2,906 iGAS episodes, 34.3% during the pandemic period were associated with acute SARS-CoV-2 effects (range by age group: 16.5-39.1%). Models incorporating cumulative SARS-CoV-2 burden showed markedly better fit (ΔAIC=−157.5); cumulative exposure was strongly associated with iGAS (IRR 1.193, 95% CI 1.151-1.235), increasing the estimated PAF to 66.7%. Cumulative effects were strongest in children (IRR 1.309). SARS-CoV-2 was comparably associated with non-invasive streptococcal disease, with no increase in invasion propensity. Cumulative streptococcal exposure was not protective (overall IRR 1.000, p=0.730); where significant, the association was positive, opposite to immunity debt predictions.

**Conclusions:** Cumulative SARS-CoV-2 burden was strongly associated with pandemic-era iGAS incidence. Cumulative streptococcal exposure did not support the immunity debt hypothesis. These ecological findings are consistent with SARS-CoV-2-associated immune dysregulation and warrant individual-level confirmation.

## INTRODUCTION

Invasive group A streptococcal (iGAS) disease is a rare but life-threatening infection with substantial morbidity and mortality, particularly among children and older adults^1,2^. Beginning in late 2022, multiple countries reported unprecedented increases in iGAS incidence, coinciding with the relaxation of COVID-19 public health measures and widespread SARS-CoV-2 transmission^3–5^. These observations have prompted debate regarding the mechanisms underlying the post-pandemic iGAS surge.

Two competing hypotheses have emerged. The “immunity debt” hypothesis^6^ proposes that reduced exposure to endemic pathogens during non-pharmaceutical interventions (NPIs) resulted in diminished population immunity, predicting that reduced prior streptococcal exposure should increase susceptibility upon pathogen reintroduction. An alternative explanation is that SARS-CoV-2 infection itself increases susceptibility to secondary bacterial infections through both acute viral-bacterial synergy and persistent immune dysregulation, particularly following repeated infections^7,8^. Unlike the immunity debt framework, this hypothesis predicts that cumulative SARS-CoV-2 exposure, rather than streptococcal deficit, should drive iGAS risk.

Previous studies have examined respiratory virus-iGAS associations, with one Dutch analysis attributing 25-34% of the iGAS surge to influenza A and RSV^9^. However, these analyses examined each pathogen independently without joint modeling or rigorous control for seasonal co-occurrence^3,4,9^, raising the possibility that apparent viral effects reflect confounding rather than independent causal contributions.

We evaluated these competing explanations using 13 years of population-based surveillance data from Ontario, Canada. We quantified acute and cumulative SARS-CoV-2 associations with iGAS incidence, assessed specificity through negative control analyses of influenza and RSV, evaluated a specific prediction of the immunity debt hypothesis using cumulative streptococcal exposure, and examined whether SARS-CoV-2 increased streptococcal susceptibility broadly or selectively enhanced invasion propensity.

## METHODS

### Study Design and Setting

We conducted a population-based ecological time-series analysis of iGAS disease in the Greater Toronto-Hamilton Area Plus (GTHA+) region of central Ontario, Canada. The GTHA+ comprises 14 Public Health Units with a combined population in 2024 of approximately 11.2 million residents, representing 27% of the Canadian population (**Supplementary Appendix 1**)^10^. The analysis period extended from March 29, 2011, through March 28, 2024 (676 weeks), including a 9-year pre-pandemic baseline (to March 14, 2020). Annual population estimates by age group and sex from Statistics Canada^10^ served as offsets in regression models.

### Case Ascertainment

Weekly iGAS counts were ascertained from two linked administrative databases maintained by the Canadian Institute for Health Information (CIHI)^11^: the Discharge Abstract Database (DAD) and the National Ambulatory Care Reporting System (NACRS). Invasive GAS was defined as sepsis due to group A streptococcus (ICD-10-CA code A40.0) in any diagnosis position, or the co-occurrence of an invasive syndrome code (meningitis G00.2, necrotizing fasciitis M72.2/M72.6, pyothorax J86.0/J86.9, pyogenic arthritis M00.0/M00.2, or endocarditis I33.0) with B95.0 (group A streptococcus as cause). Cross-database deduplication using encrypted identifiers ensured one episode per person. Non-invasive streptococcal disease and total streptococcal disease were ascertained using analogous methods (Supplementary Appendix 7). Weekly case counts were aggregated by age group (0-19, 20-64, ≥65 years) and sex, yielding 6 age-sex strata per week.

### Respiratory Virus Exposures

SARS-CoV-2 exposure during pandemic period 1 (March 2020-August 2022) was based on test-adjusted reported case counts from Ontario’s Case and Contact Management System (CCM) ^12^ from March 2020 to August 2022 (using methodology described in Fisman et al. ^13^ and Bosco et al. ^14^). From September 2022 onward (pandemic period 2), and for all other viruses throughout, we used percent positivity from the Respiratory Virus Detection Surveillance System (RVDSS)^15^. The two periods also differ in public health context: period 1 was characterized by active non-pharmaceutical interventions aimed at slowing spread of SARS-CoV-2^14,16,17^, while period 2 followed their relaxation. All viral measures were normalized to standard deviation units. Cumulative SARS-CoV-2 burden was constructed as the running sum of standardized weekly exposure from March 2020, representing accumulated population-level viral exposure over time. Acute effects were modeled using a 2-week lag based on the expected interval between respiratory viral infection and secondary bacterial disease^18–21^.

### Statistical Analysis

Weekly iGAS counts were modeled using negative binomial panel regression (6 age-sex strata) with population offset, Fourier seasonal terms (annual periodicity), quadratic time trends, and age-sex fixed effects. The primary analysis examined acute SARS-CoV-2 associations. In exploratory analyses, cumulative SARS-CoV-2 burden was added to assess progressive effects independent of acute exposure; model fit was compared using AIC. Identical structures were applied to influenza and RSV as negative controls. The immunity debt hypothesis^6^ predicts that cumulative streptococcal exposure should be inversely associated with iGAS risk (IRR<1); we tested this specific prediction by including cumulative streptococcal burden in period 2 models. We illustrate these competing frameworks and their empirical predictions in **Supplementary Appendix 2**.

To distinguish increased susceptibility from increased invasion propensity, we replaced the population offset with total streptococcal disease. PAFs were estimated by counterfactual prediction^22^, setting SARS-CoV-2 coefficients to zero.

### Ethical Approval

This study used de-identified administrative data in the CIHI Secure Access Environment and was approved by the Research Ethics Board of the University of Toronto (Protocol # 41690).

## RESULTS

### Temporal Trends

Over 13 years, we identified 2,906 iGAS episodes, yielding a crude incidence of 2.23 per 100,000 person-years (95% CI 2.13-2.34) (**Table 1**, **Figure 1**). Incidence declined during pandemic period 1 to 1.49 per 100,000 (IRR 0.70, 95% CI 0.62-0.79 vs. pre-pandemic), then rose during period 2 to 3.10 per 100,000 (IRR 2.03, 95% CI 1.78-2.33). Adults aged ≥65 had the highest iGAS risk throughout (IRR 2.85 vs. ages 20-64) and males had consistently elevated risk (IRR 1.33). Children’s relative risk shifted markedly: from 40% below adults pre-pandemic (IRR 0.61), to 83% below during period 1 (IRR 0.17), to parity during period 2 (IRR 1.11). In absolute terms, pediatric iGAS incidence fell from 1.01 per 100,000 pre-pandemic to 0.24 per 100,000 during period 1, before rebounding to 2.48 per 100,000 during period 2, a rate 2.5-fold above the pre-pandemic baseline and exceeding the pre-pandemic rate in working-age adults (2.22 per 100,000).

**Figure 1.**
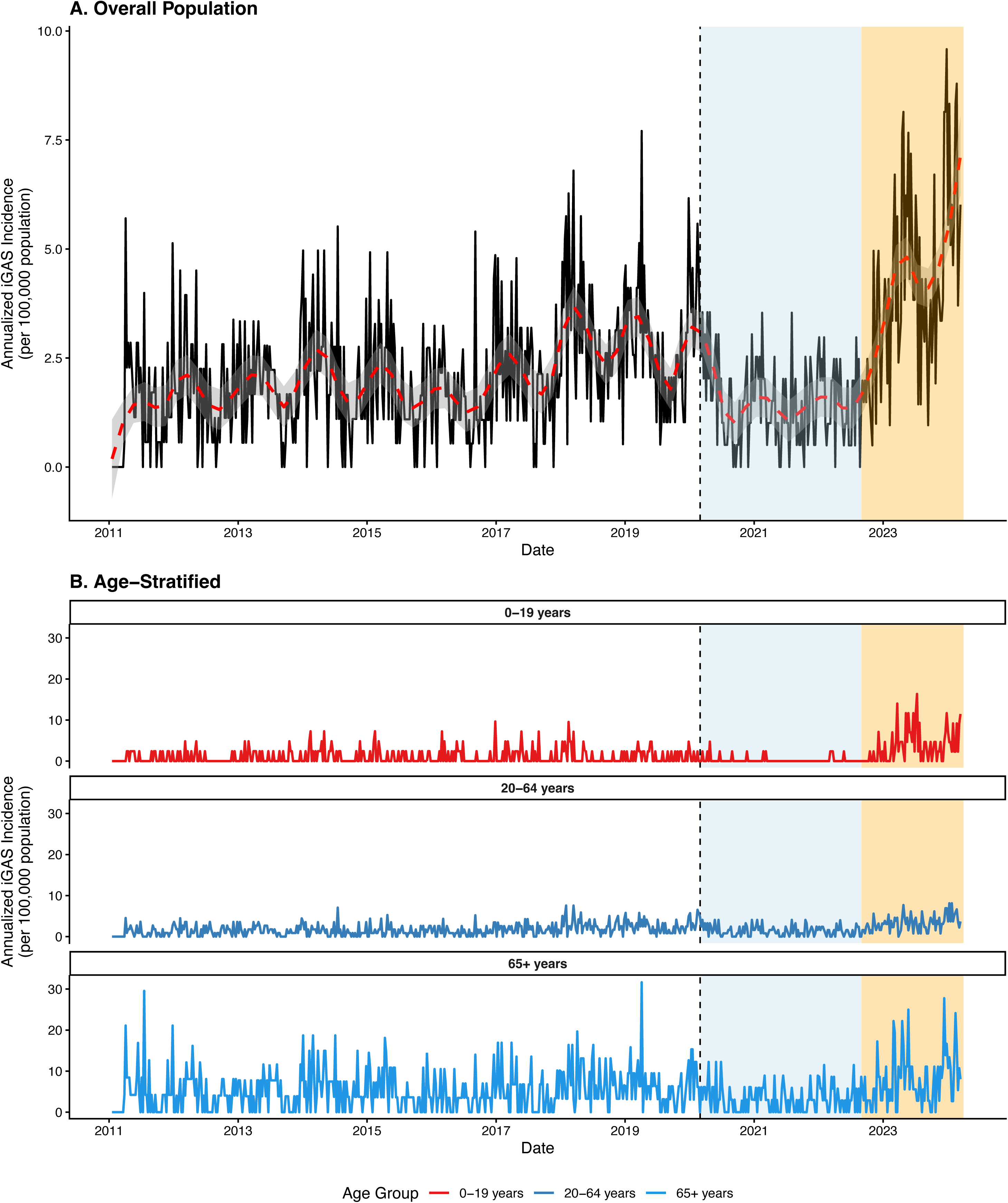
Temporal Trends in Invasive Group A Streptococcal Disease Incidence. (A) Weekly annualized iGAS incidence (per 100,000 population) in the GTHA+ region, March 2011-March 2024. Black line, observed rates; dashed red line, fitted seasonal trend. Blue shading, pandemic period 1 (March 2020-August 2022); orange shading, pandemic period 2 (September 2022-March 2024). (B) Age-stratified incidence for children (0-19 years), working-age adults (20-64 years), and older adults (≥65 years).

**Table 1.**
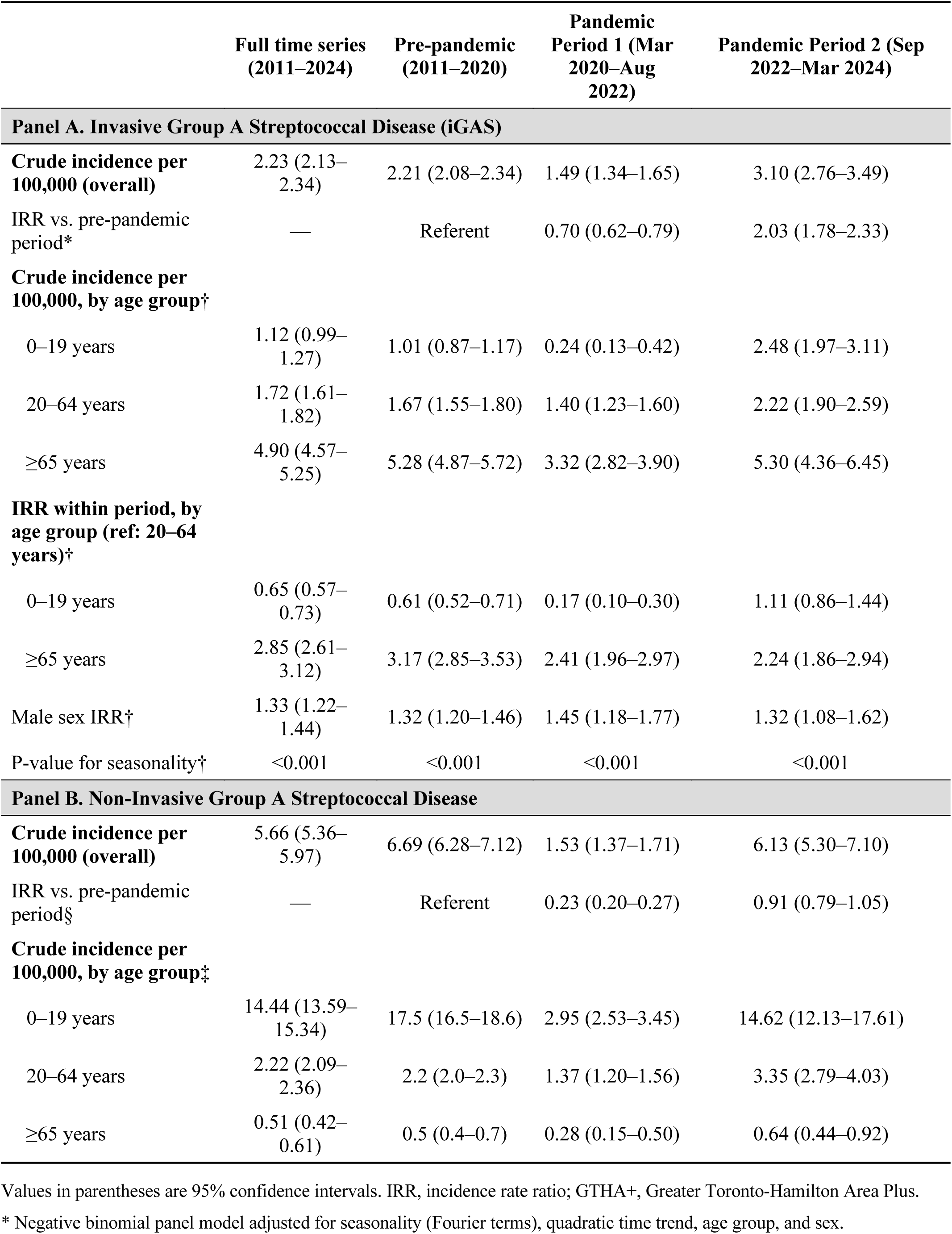
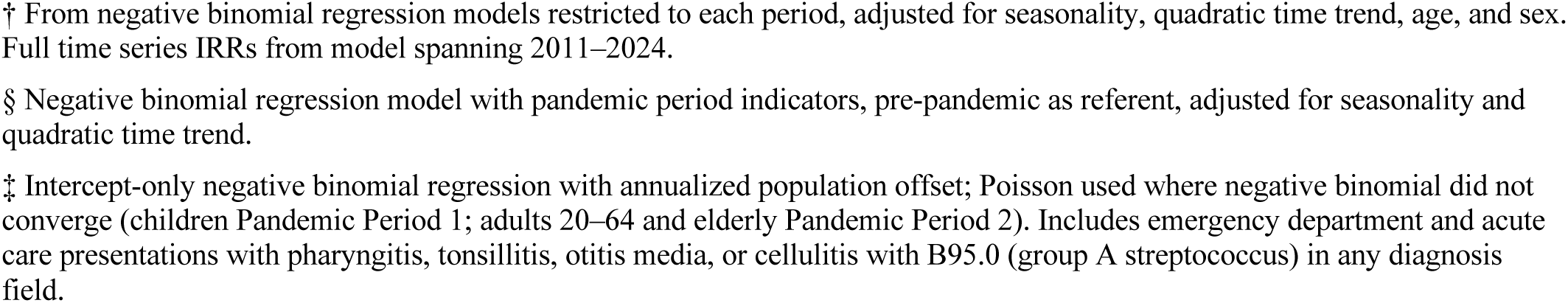
Invasive and Non-Invasive Group A Streptococcal Disease Incidence and Risk Factors by Study Period, GTHA+, 2011–2024.

|  | Full time series<br>(2011–2024) | Pre-pandemic<br>(2011–2020) | Pandemic<br>Period 1 (Mar<br>2020–Aug<br>2022) | Pandemic Period 2 (Sep<br>2022–Mar 2024) |
| --- | --- | --- | --- | --- |
| **Panel A. Invasive Group A Streptococcal Disease (iGAS)** |  |  |  |  |
| **Crude incidence per 100,000 (overall)** | 2.23 (2.13–2.34) | 2.21 (2.08–2.34) | 1.49 (1.34–1.65) | 3.10 (2.76–3.49) |
| IRR vs. pre-pandemic period* | — | Referent | 0.70 (0.62–0.79) | 2.03 (1.78–2.33) |
| **Crude incidence per 100,000, by age group†** |  |  |  |  |
| 0–19 years | 1.12 (0.99–1.27) | 1.01 (0.87–1.17) | 0.24 (0.13–0.42) | 2.48 (1.97–3.11) |
| 20–64 years | 1.72 (1.61–1.82) | 1.67 (1.55–1.80) | 1.40 (1.23–1.60) | 2.22 (1.90–2.59) |
| ≥65 years | 4.90 (4.57–5.25) | 5.28 (4.87–5.72) | 3.32 (2.82–3.90) | 5.30 (4.36–6.45) |
| **IRR within period, by age group (ref: 20–64 years)†** |  |  |  |  |
| 0–19 years | 0.65 (0.57–0.73) | 0.61 (0.52–0.71) | 0.17 (0.10–0.30) | 1.11 (0.86–1.44) |
| ≥65 years | 2.85 (2.61–3.12) | 3.17 (2.85–3.53) | 2.41 (1.96–2.97) | 2.24 (1.86–2.94) |
| Male sex IRR† | 1.33 (1.22–1.44) | 1.32 (1.20–1.46) | 1.45 (1.18–1.77) | 1.32 (1.08–1.62) |
| P-value for seasonality† | <0.001 | <0.001 | <0.001 | <0.001 |
| **Panel B. Non-Invasive Group A Streptococcal Disease** |  |  |  |  |
| **Crude incidence per 100,000 (overall)** | 5.66 (5.36–5.97) | 6.69 (6.28–7.12) | 1.53 (1.37–1.71) | 6.13 (5.30–7.10) |
| IRR vs. pre-pandemic period§ | — | Referent | 0.23 (0.20–0.27) | 0.91 (0.79–1.05) |
| **Crude incidence per 100,000, by age group‡** |  |  |  |  |
| 0–19 years | 14.44 (13.59–15.34) | 17.5 (16.5–18.6) | 2.95 (2.53–3.45) | 14.62 (12.13–17.61) |
| 20–64 years | 2.22 (2.09–2.36) | 2.2 (2.0–2.3) | 1.37 (1.20–1.56) | 3.35 (2.79–4.03) |
| ≥65 years | 0.51 (0.42–0.61) | 0.5 (0.4–0.7) | 0.28 (0.15–0.50) | 0.64 (0.44–0.92) |

Values in parentheses are 95% confidence intervals. IRR, incidence rate ratio; GTHA+, Greater Toronto-Hamilton Area Plus.
\* Negative binomial panel model adjusted for seasonality (Fourier terms), quadratic time trend, age group, and sex.
† From negative binomial regression models restricted to each period, adjusted for seasonality, quadratic time trend, age, and sex. Full time series IRRs from model spanning 2011–2024.
§ Negative binomial regression model with pandemic period indicators, pre-pandemic as referent, adjusted for seasonality and quadratic time trend.
‡ Intercept-only negative binomial regression with annualized population offset; Poisson used where negative binomial did not converge (children Pandemic Period 1; adults 20–64 and elderly Pandemic Period 2). Includes emergency department and acute care presentations with pharyngitis, tonsillitis, otitis media, or cellulitis with B95.0 (group A streptococcus) in any diagnosis field.

Pre-pandemic non-invasive streptococcal disease incidence was highest among children aged 0-19 years (17.5 per 100,000 per year; 95% CI 16.5-18.6), approximately 8-fold higher than in working-age adults (2.2 per 100,000) and 35-fold higher than in older adults (0.5 per 100,000) (**Table 1**). During pandemic period 1, non-invasive streptococcal incidence declined markedly across all age groups (IRR 0.23, 95% CI 0.20-0.27), before rebounding toward pre-pandemic levels during pandemic period 2 (IRR 0.91, 95% CI 0.79-1.05).

### SARS-CoV-2 Associations

Each 1-SD increase in acute SARS-CoV-2 exposure was associated with a 14% increase in iGAS during period 1 (IRR 1.136, 95% CI 1.020-1.264, p=0.020) and 38% during period 2 (IRR 1.384, 95% CI 1.172-1.635, p<0.001). Acute effects were statistically significant in working-age adults during both periods and in older adults during period 2, but were not significant in children during either period, likely reflecting the smaller number of pediatric cases (**Table 2, Panel A**).

**Table 2.**
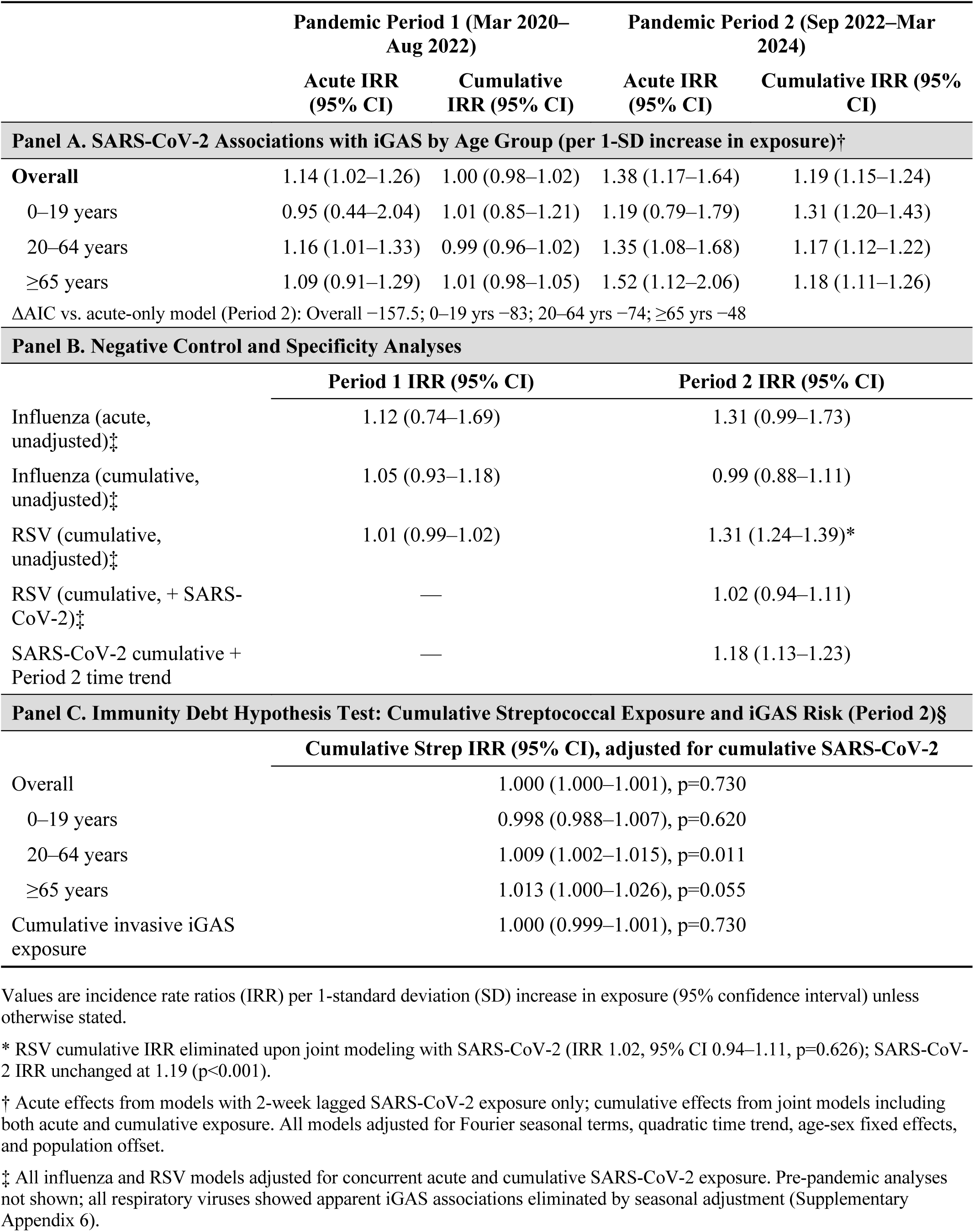
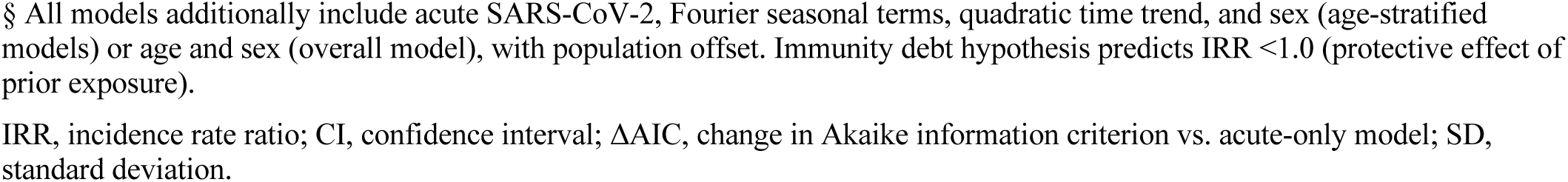
SARS-CoV-2 Associations with Invasive Group A Streptococcal Disease, Negative Control Analyses, and Immunity Debt Hypothesis Tests.

|  | Pandemic Period 1 (Mar 2020–<br>Aug 2022) |  | Pandemic Period 2 (Sep 2022–Mar<br>2024) |  |
| --- | --- | --- | --- | --- |
|  | Acute IRR<br>(95% CI) | Cumulative<br>IRR (95% CI) | Acute IRR<br>(95% CI) | Cumulative IRR (95%<br>CI) |
| Panel A. SARS-CoV-2 Associations with iGAS by Age Group (per 1-SD increase in exposure)† |  |  |  |  |
| Overall | 1.14 (1.02–1.26) | 1.00 (0.98–1.02) | 1.38 (1.17–1.64) | 1.19 (1.15–1.24) |
| 0–19 years | 0.95 (0.44–2.04) | 1.01 (0.85–1.21) | 1.19 (0.79–1.79) | 1.31 (1.20–1.43) |
| 20–64 years | 1.16 (1.01–1.33) | 0.99 (0.96–1.02) | 1.35 (1.08–1.68) | 1.17 (1.12–1.22) |
| ≥65 years | 1.09 (0.91–1.29) | 1.01 (0.98–1.05) | 1.52 (1.12–2.06) | 1.18 (1.11–1.26) |
| ΔAIC vs. acute-only model (Period 2): Overall –157.5; 0–19 yrs –83; 20–64 yrs –74; ≥65 yrs –48 |  |  |  |  |
| Panel B. Negative Control and Specificity Analyses |  |  |  |  |
|  | Period 1 IRR (95% CI) |  | Period 2 IRR (95% CI) |  |
| Influenza (acute,<br>unadjusted)‡ | 1.12 (0.74–1.69) |  | 1.31 (0.99–1.73) |  |
| Influenza (cumulative,<br>unadjusted)‡ | 1.05 (0.93–1.18) |  | 0.99 (0.88–1.11) |  |
| RSV (cumulative,<br>unadjusted)‡ | 1.01 (0.99–1.02) |  | 1.31 (1.24–1.39)* |  |
| RSV (cumulative, + SARS-CoV-2)‡ | — |  | 1.02 (0.94–1.11) |  |
| SARS-CoV-2 cumulative +<br>Period 2 time trend | — |  | 1.18 (1.13–1.23) |  |
| Panel C. Immunity Debt Hypothesis Test: Cumulative Streptococcal Exposure and iGAS Risk (Period 2)§ |  |  |  |  |
|  | Cumulative Strep IRR (95% CI), adjusted for cumulative SARS-CoV-2 |  |  |  |
| Overall | 1.000 (1.000–1.001), p=0.730 |  |  |  |
| 0–19 years | 0.998 (0.988–1.007), p=0.620 |  |  |  |
| 20–64 years | 1.009 (1.002–1.015), p=0.011 |  |  |  |
| ≥65 years | 1.013 (1.000–1.026), p=0.055 |  |  |  |
| Cumulative invasive iGAS<br>exposure | 1.000 (0.999–1.001), p=0.730 |  |  |  |

Values are incidence rate ratios (IRR) per 1-standard deviation (SD) increase in exposure (95% confidence interval) unless otherwise stated.
\* RSV cumulative IRR eliminated upon joint modeling with SARS-CoV-2 (IRR 1.02, 95% CI 0.94–1.11, p=0.626); SARS-CoV-2 IRR unchanged at 1.19 (p<0.001).
† Acute effects from models with 2-week lagged SARS-CoV-2 exposure only; cumulative effects from joint models including both acute and cumulative exposure. All models adjusted for Fourier seasonal terms, quadratic time trend, age-sex fixed effects, and population offset.
‡ All influenza and RSV models adjusted for concurrent acute and cumulative SARS-CoV-2 exposure. Pre-pandemic analyses not shown; all respiratory viruses showed apparent iGAS associations eliminated by seasonal adjustment (Supplementary Appendix 6).
§ All models additionally include acute SARS-CoV-2, Fourier seasonal terms, quadratic time trend, and sex (age-stratified models) or age and sex (overall model), with population offset. Immunity debt hypothesis predicts IRR <1.0 (protective effect of prior exposure).
IRR, incidence rate ratio; CI, confidence interval; ΔAIC, change in Akaike information criterion vs. acute-only model; SD, standard deviation.

In exploratory analyses, cumulative SARS-CoV-2 burden showed no association during period 1 (IRR 0.998, p=0.824) but was strongly associated with iGAS during period 2 (IRR 1.193, 95% CI 1.151-1.235, p<0.001; ΔAIC=−157.5). The improvement in model fit with cumulative burden was substantial and consistent across all age groups (ΔAIC: −83 for children, −74 for working-age adults, −48 for older adults), indicating that cumulative exposure captured meaningful variation in iGAS risk beyond what acute exposure alone explained. Strikingly, while children showed the weakest acute effects, they demonstrated the strongest cumulative association (IRR 1.309, 95% CI 1.198-1.430) (**Figure 2**, **Table 2** **Panel A**).

**Figure 2.**
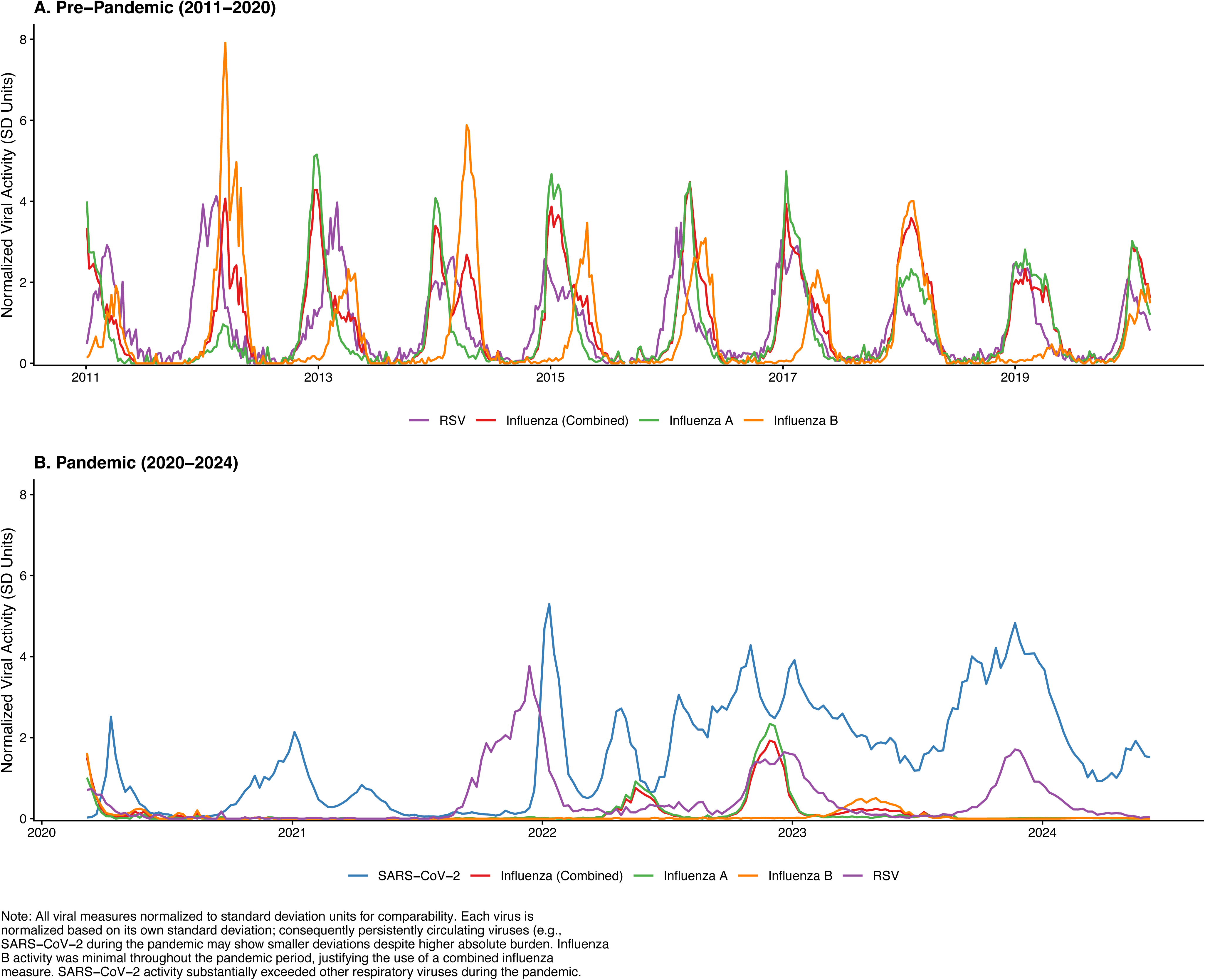
Acute and Cumulative SARS-CoV-2 Associations with iGAS by Age Group and Pandemic Period. (A) Incidence rate ratios per 1-SD increase in 2-week lagged SARS-CoV-2 exposure, from acute-only models. (B) Cumulative SARS-CoV-2 IRRs from joint models. All models adjusted for seasonal harmonics, time trends, and sex.

Using counterfactual prediction, 34.3% of pandemic-era iGAS cases were associated with acute SARS-CoV-2 effects (**Figure 3**). Acute PAFs were highest among older adults (39.1%) and lowest among children (16.5%). Incorporating cumulative burden increased the overall estimate to 66.7%, with the largest increases in children (98.5%) and older adults (96.1%); these exploratory cumulative PAFs should be interpreted cautiously given uncertainty in the cumulative exposure measure.

**Figure 3.**
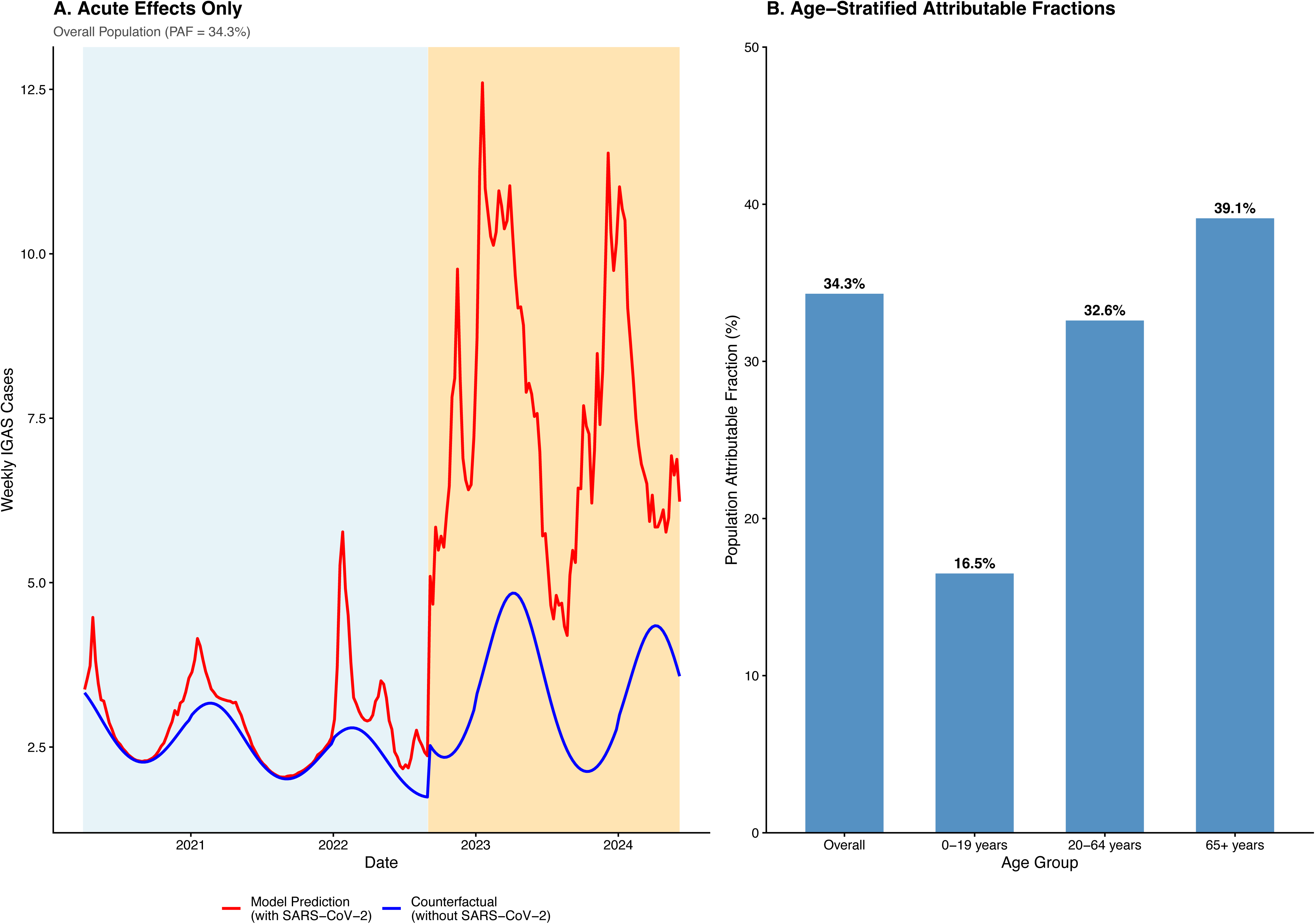
Population Attributable Fractions for Acute SARS-CoV-2 Effects. (A) Counterfactual prediction for the overall population. Red line, model prediction; blue line, counterfactual without SARS-CoV-2 (PAF=34.3%). (B) Age-stratified acute PAFs.

### Negative Controls and Specificity

In the pre-pandemic period, all respiratory viruses showed apparent associations with iGAS that were entirely explained by seasonal confounding: without Fourier harmonic adjustment, IRRs for influenza, RSV, influenza A, and influenza B ranged from 1.11 to 1.21 (all p<0.001); after adjustment, all became null (all p>0.13) (**Supplementary** **Appendix 6**). This finding indicates that previously reported virus-iGAS associations lacking adequate seasonal adjustment likely reflect shared winter seasonality rather than independent causal effects.

During the pandemic, acute and cumulative influenza were null in both periods (acute period 2 IRR 1.31, 95% CI 0.99-1.73; cumulative IRR 0.99, 95% CI 0.88-1.11) (**Table 2, Panel B**). Cumulative RSV was associated with iGAS when modeled alone (IRR 1.31, 95% CI 1.24-1.39, p<0.001) but this association was fully eliminated in joint models with SARS-CoV-2 (RSV IRR 1.02, 95% CI 0.94-1.11, p=0.626; SARS-CoV-2 IRR unchanged at 1.19, p<0.001), indicating that the apparent RSV-iGAS association reflected confounding by co-circulating SARS-CoV-2 rather than an independent RSV effect. The SARS-CoV-2 cumulative association was also robust to inclusion of a period 2-specific linear time trend, which was itself non-significant (time trend IRR 1.14, 95% CI 0.97-1.32, p=0.108; SARS-CoV-2 IRR 1.18, 95% CI 1.13-1.23, p<0.001), indicating that the association is not explained by a general secular increase in iGAS risk during this period.

### Non-Invasive Streptococcal Disease and Invasion Propensity

Cumulative SARS-CoV-2 burden was comparably associated with non-invasive streptococcal incidence (IRR 1.216, 95% CI 1.184-1.249) and iGAS (IRR 1.193, 95% CI 1.151-1.235) during period 2 (**Supplementary** **Appendix 7**). The near-identical effect sizes across invasive and non-invasive outcomes indicate that SARS-CoV-2 increased susceptibility to streptococcal disease broadly, affecting the full spectrum from superficial infection to life-threatening invasive disease. Consistent with this interpretation, when total streptococcal disease replaced population as the model offset, thereby estimating iGAS risk conditional on streptococcal infection, SARS-CoV-2 showed no significant association with invasion propensity during period 2 (cumulative IRR 1.037, p=0.101). This pattern points toward a mechanism operating at the level of susceptibility to streptococcal acquisition, rather than at the step of mucosal barrier breach or tissue invasion.

### Evaluation of the Immunity Debt Hypothesis

The immunity debt hypothesis predicts that cumulative streptococcal exposure should be inversely associated with iGAS risk (IRR<1.0), reflecting the protective effect of prior immune priming. In joint models including both cumulative SARS-CoV-2 and cumulative streptococcal burden, this prediction was not supported in any age group (**Table 2, Panel C**). Cumulative SARS-CoV-2 remained strongly associated with iGAS across all age groups (IRRs 1.14-1.31, all p<0.001) with no attenuation upon inclusion of streptococcal burden. Cumulative streptococcal exposure showed no protective association overall (IRR 1.000, 95% CI 1.000-1.001, p=0.730) or among children (IRR 0.998, 95% CI 0.988-1.007, p=0.620), the age group most often cited in immunity debt arguments. Among working-age adults, cumulative streptococcal burden showed a modest positive association (IRR 1.009, 95% CI 1.002-1.015, p=0.011), a direction opposite to that predicted by immunity debt and consistent instead with greater streptococcal circulation providing more opportunities for invasion. Among older adults the association was null (IRR 1.013, 95% CI 1.000-1.026, p=0.055). Across all six formulations tested, three streptococcal exposure measures (invasive, non-invasive, total) in two pandemic periods, no protective effect of prior streptococcal exposure was observed.

## DISCUSSION

In this 13-year population-based study encompassing approximately 11 million residents of Ontario, cumulative SARS-CoV-2 burden was strongly associated with pandemic-era iGAS incidence and substantially improved model fit beyond acute effects. Acute SARS-CoV-2 effects accounted for an estimated 34.3% of pandemic-era cases, with exploratory cumulative models suggesting up to 66.7%. These associations were specific to SARS-CoV-2, robust to negative control analyses, and not explained by secular time trends. Cumulative streptococcal exposure did not support a key prediction of the immunity debt hypothesis.

The pattern of results is consistent with two mechanisms operating in concert. First, acute viral-bacterial synergy: respiratory viral infections predispose to secondary bacterial disease through impaired innate immune defenses, including neutrophil degranulation dysfunction, lymphopenia, and suppression of type I interferon signaling ^23–26^. Unlike respiratory pathogens such as pneumococcus and meningococcus, for which mucosal injury is a plausible portal-of-entry mechanism, iGAS is predominantly a skin and soft tissue pathogen, making systemic innate immune suppression a more parsimonious acute mechanism for our finding that acute SARS-CoV-2 exposure was associated with iGAS across both pandemic periods. Second, the emergence of cumulative effects during period 2, when seroprevalence of SARS-CoV-2 anti-N antibody (signifying infection rather than immunization) exceeded 60%^27^ and repeated infections were common, is consistent with reports of persistent T-cell exhaustion^7^, increased infection susceptibility following COVID-19^28^, and progressive immune alterations with repeated SARS-CoV-2 exposure. However, the cumulative exposure variable is a population-level proxy that increases monotonically with time; while the association survived time trend adjustment and negative control comparisons, causal interpretation requires individual-level confirmation.

Analyses of non-invasive streptococcal disease help characterize the nature of the association. SARS-CoV-2 was comparably associated with non-invasive and invasive streptococcal disease, and showed no association with invasion propensity when streptococcal disease burden was used as the offset. This pattern suggests increased susceptibility to streptococcal infection broadly, rather than selective enhancement of mechanisms enabling tissue invasion.

Our results do not support a specific, testable prediction of the immunity debt hypothesis^6,29^: that reduced prior streptococcal exposure should increase iGAS risk. Cumulative streptococcal exposure was null overall and among children, the population most often emphasized in immunity debt arguments. Where significant (working-age adults), the association was positive, consistent with greater streptococcal circulation providing more opportunities for invasion rather than reduced exposure conferring vulnerability. We acknowledge that immunity debt encompasses broader concepts including cross-pathogen immunity and cohort-specific exposure gaps; our test addresses one falsifiable prediction using available data.

The negative control analyses strengthen the case for SARS-CoV-2 specificity. Pre-pandemic analyses demonstrated that all respiratory viruses showed apparent iGAS associations entirely explained by seasonal confounding, a finding with implications for previous studies lacking adequate seasonal adjustment^9^. During the pandemic, the cumulative RSV-iGAS association was eliminated by joint modeling with SARS-CoV-2, indicating confounding by co-circulation rather than an independent RSV effect.

The pediatric pattern we observed warrants emphasis. Children had the lowest acute PAF (16.5%) but the strongest cumulative association (IRR 1.309) and the most dramatic shift in relative iGAS risk across periods. This pattern is difficult to reconcile with immunity debt, which predicts the largest rebounds in children due to missed exposures, but is consistent with cumulative effects from repeated SARS-CoV-2 infections.

Key limitations include the ecological design, which precludes individual-level causal inference; the change in SARS-CoV-2 exposure metrics between periods; and the inherent correlation between cumulative exposure and time, which sensitivity analyses can mitigate but not fully resolve. PAF estimates, particularly the exploratory cumulative estimates, lack formal uncertainty bounds and should be interpreted as indicative of magnitude rather than precise.

Non-invasive streptococcal disease was ascertained from emergency department visits and select ambulatory care settings reporting to CIHI, and does not capture the majority of primary care encounters managed in family physician and walk-in clinic settings; shifts in care-seeking behaviour across pandemic periods may therefore affect period-to-period comparisons of non-invasive disease incidence and should be considered when interpreting apparent differences in the ratio of invasive to non-invasive streptococcal presentations. Our data derive from a single Canadian region, though iGAS patterns mirror global reports^3,5,30^. Strengths include the 9-year baseline enabling seasonal confounding demonstration, the large population supporting age-stratified analyses, and multiple complementary approaches testing competing hypotheses.

These findings do not exclude some contribution of reduced pathogen exposure during NPIs to broader infection patterns. Rather, they indicate that this mechanism is insufficient to explain the iGAS surge. If these associations reflect causal effects, reducing cumulative SARS-CoV-2 exposure, particularly in children, could reduce severe bacterial disease burden.

Individual-level studies with immunological endpoints are needed to confirm whether the population-level patterns observed here reflect SARS-CoV-2-mediated immune dysregulation.

## CONCLUSIONS

In this 13-year population-based study, cumulative SARS-CoV-2 burden was strongly and specifically associated with pandemic-era iGAS incidence. SARS-CoV-2 was associated with increased streptococcal disease broadly without evidence of selectively increasing invasion propensity. Evaluation of the impact of cumulative streptococcal exposure did not support the immunity debt hypothesis. These ecological findings are consistent with SARS-CoV-2-associated immune dysregulation as a contributor to the post-pandemic iGAS surge and warrant confirmation through individual-level studies.

## Data Availability

Code Availability: DOI: 10.5281/zenodo.19098309
Data Availability: Individual-level administrative health data were accessed within the
Canadian Institute for Health Information Secure Access Environment. Respiratory virus
surveillance data are publicly available through Public Health Agency of Canada.

https://github.com/fismanda/igas-pandemic-ontario

## ACKNOWLEDGEMENTS

The authors thank the Canadian Institute for Health Information (CIHI) for providing access to the Discharge Abstract Database and National Ambulatory Care Reporting System through the Secure Access Environment, and the Public Health Agency of Canada for the Respiratory Virus Detection Surveillance System data. The analyses, conclusions, opinions, and statements expressed herein are solely those of the authors and do not reflect those of the data sources; no endorsement is intended or should be inferred.

## AI DISCLOSURE

AI-assisted technologies (Claude, Anthropic and ChatGPT, OpenAI) were used during manuscript preparation for statistical code review and editorial revision. All AI-generated content was reviewed, verified, and edited by the authors, who take full responsibility for the accuracy and integrity of the work.

## Code Availability

DOI: 10.5281/zenodo.19098309

## Data Availability

Individual-level administrative health data were accessed within the Canadian Institute for Health Information Secure Access Environment. Respiratory virus surveillance data are publicly available through Public Health Agency of Canada.

## Supplementary Appendix 1: Study Region Definition and Population Characteristics

### Geographic Coverage

The Greater Toronto-Hamilton Area Plus (GTHA+) comprises 14 Public Health Units (PHUs) in central Ontario, Canada (Figure S1). The region consists of the 6 PHUs of the Greater Toronto-Hamilton Area (GTHA): Toronto, Hamilton, Peel, York, Durham, and Halton; plus 8 bordering PHUs: Simcoe-Muskoka, Haliburton Kawartha Pineridge, Peterborough, Wellington-Dufferin-Guelph, Waterloo, Brant, Niagara, and Haldimand-Norfolk.

### Rationale for Study Region

The GTHA+ was selected to provide a large population concentrated in a small geographic area, facilitating exposure assessment in ecological analyses. The region represents 67% of Ontario’s population and approximately 27% of Canada’s population, while comprising only 5% of Ontario’s total land area. This geographic concentration provides assurance that relevant exposures (viral activity, environmental covariates) have been experienced by most of the study population. The relative abundance of healthcare institutions in this region also provides advantages in terms of statistical power for health system-derived outcomes.

### Population Characteristics

Population estimates for each PHU were obtained from Statistics Canada population tables,^1^ which provide annual estimates for both census and non-census years. As of 2024, the GTHA+ had an estimated population of 11.2 million residents. The region experienced substantial growth during the study period, with overall population increasing 41.3% between 2001 and 2024 (**Figure S2**). Growth rates varied across constituent PHUs, ranging from 18.6% (Haldimand-Norfolk) to 61.1% (Peel).

**Figure S1.**
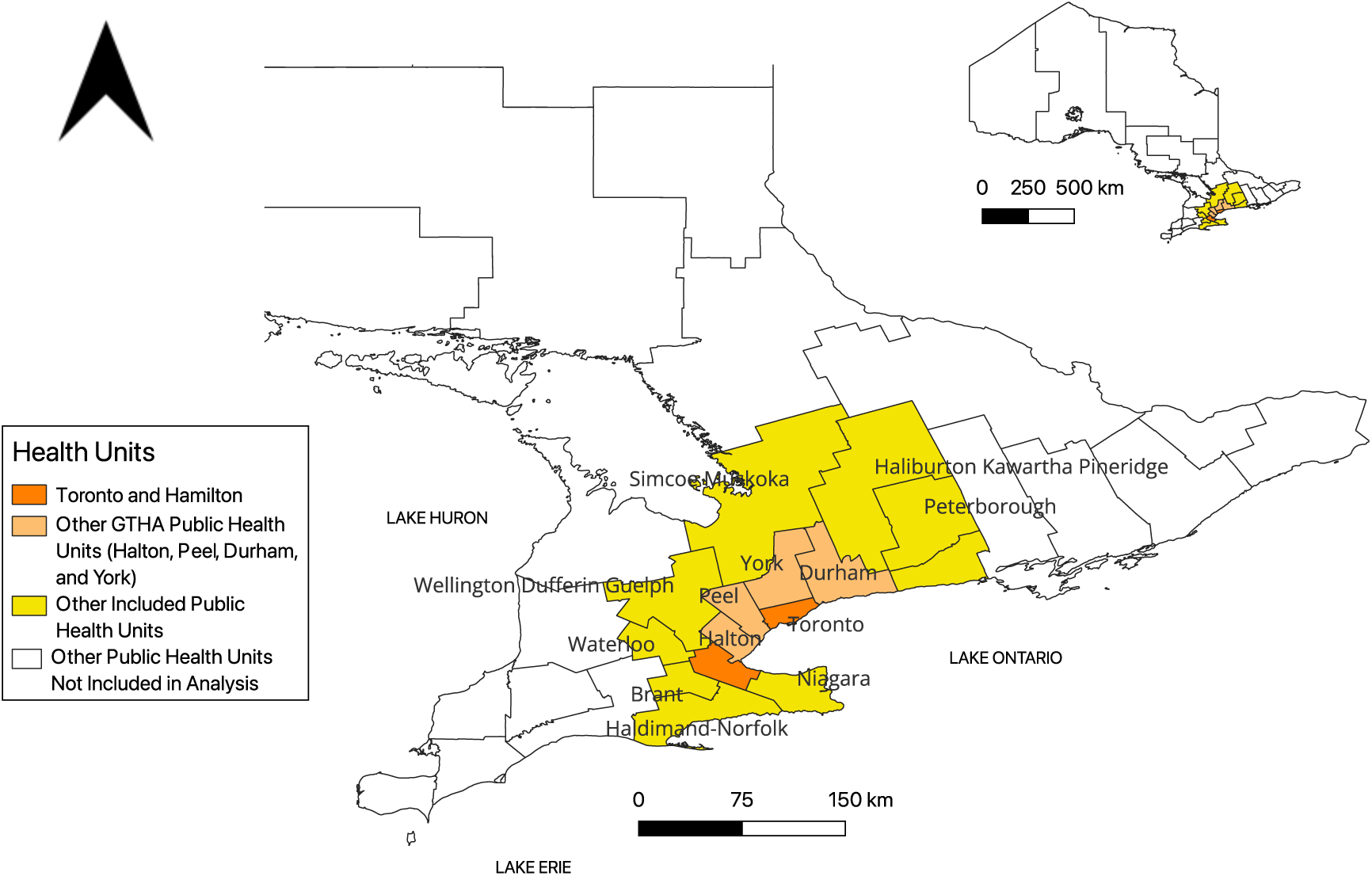
Map of Public Health Units included in the GTHA+ study region. The main map shows the 14 Public Health Units comprising the GTHA+ region within southern Ontario. Toronto and Hamilton (core GTHA cities) are shown in dark orange; other GTHA Public Health Units (Halton, Peel, Durham, and York) are shown in light orange; and the 8 additional included PHUs (Simcoe-Muskoka, Haliburton Kawartha Pineridge, Peterborough, Wellington-Dufferin-Guelph, Waterloo, Brant, Niagara, and Haldimand-Norfolk) are shown in yellow. Public Health Units not included in the analysis are shown in white. The inset map (upper right) shows the location of the GTHA+ region (yellow) within the province of Ontario, Canada. Scale bars indicate distances of 0-150 km for the main map and 0-500 km for the inset.

**Figure S2.**
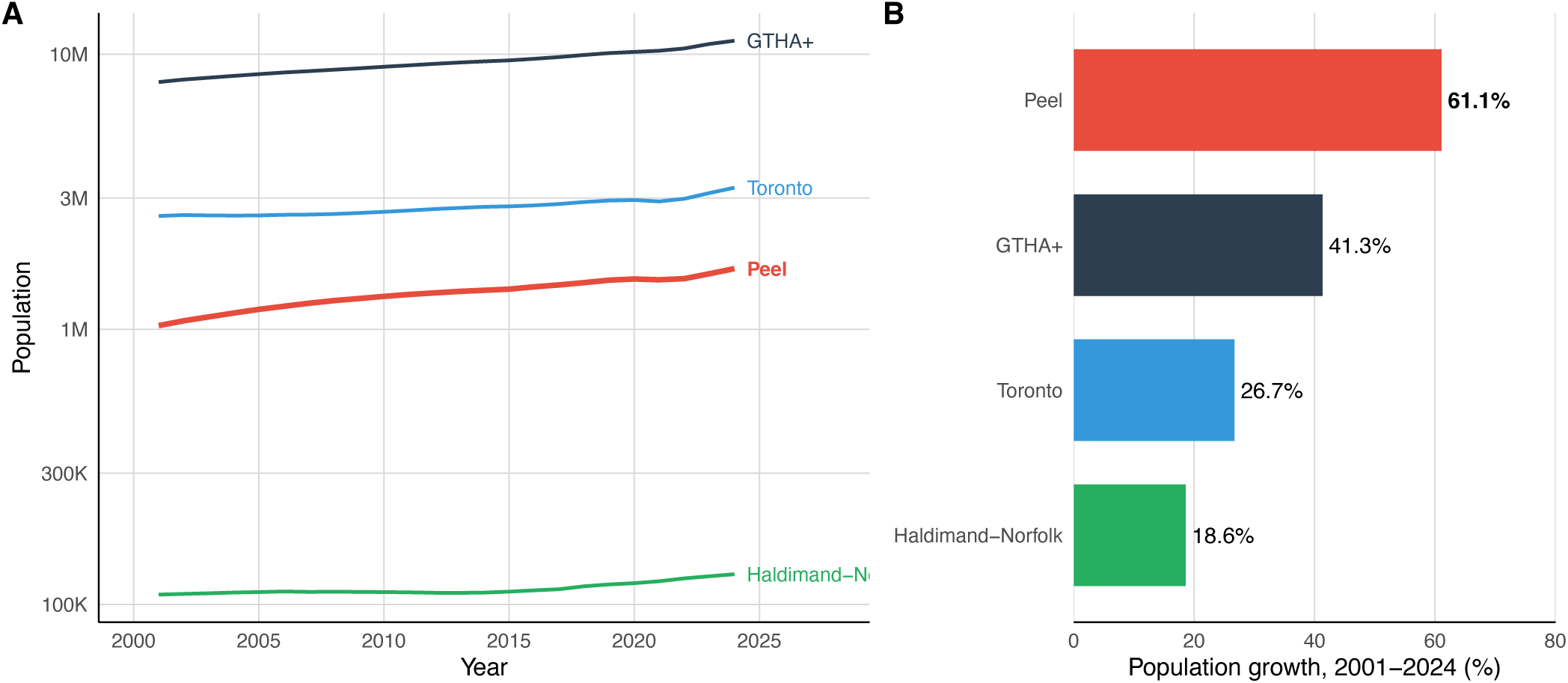
Population growth in the GTHA+ region, 2001-2024. Panel A shows population trajectories on a log scale for the overall GTHA+ region and for the largest (Toronto), smallest (Haldimand-Norfolk), and most rapidly growing (Peel) PHU. Panel B shows percent population growth over the study period for these PHUs.

## Supplementary Appendix 2: Competing Mechanistic Hypotheses for the Post-Pandemic Invasive Group A Streptococcal Disease Surge

Three mechanistic hypotheses have been proposed to explain the post-pandemic surge in invasive group A streptococcal (iGAS) disease. **Figure S2** illustrates the causal pathways, predicted temporal patterns, and empirical consistency of each hypothesis.

The **immunity debt hypothesis** proposes that non-pharmaceutical interventions (NPIs) reduced population exposure to group A streptococcus, causing immunity to wane. Upon NPI relaxation and pathogen reintroduction, a surge in susceptibles would produce a discrete rebound in iGAS incidence followed by return to baseline. This hypothesis predicts that cumulative streptococcal deficit should be associated with iGAS risk, that effects should be non-specific to any single pathogen, and that children, presumed to have the least prior exposure, should be most affected.

The **acute viral facilitation** hypothesis proposes that SARS-CoV-2 infection causes transient impairment of innate host defenses, including neutrophil degranulation dysfunction, lymphopenia, and suppression of type I interferon signaling, thus creating a vulnerability window for secondary bacterial infection. This hypothesis predicts that iGAS incidence should track individual COVID-19 waves with a short lag, and that the association should be specific to SARS-CoV-2 only insofar as it was the dominant respiratory virus during the pandemic period.

The **cumulative immune dysregulation hypothesis** proposes that repeated SARS-CoV-2 infections cause progressive T-cell exhaustion and immune dysfunction, leading to escalating susceptibility to bacterial disease over time. This hypothesis predicts a progressive elevation in iGAS baseline, distinct from wave-mirroring, a strong dose-response relationship with cumulative SARS-CoV-2 burden, and the strongest effects in individuals with the highest cumulative infection burden.

Panel E of Figure S2 summarizes the consistency of each hypothesis with empirical tests conducted in this study. The cumulative immune dysregulation hypothesis was supported by all five applicable tests, while immunity debt was supported by none of four applicable tests, and acute facilitation by both of its two applicable tests.

**Figure S3.**
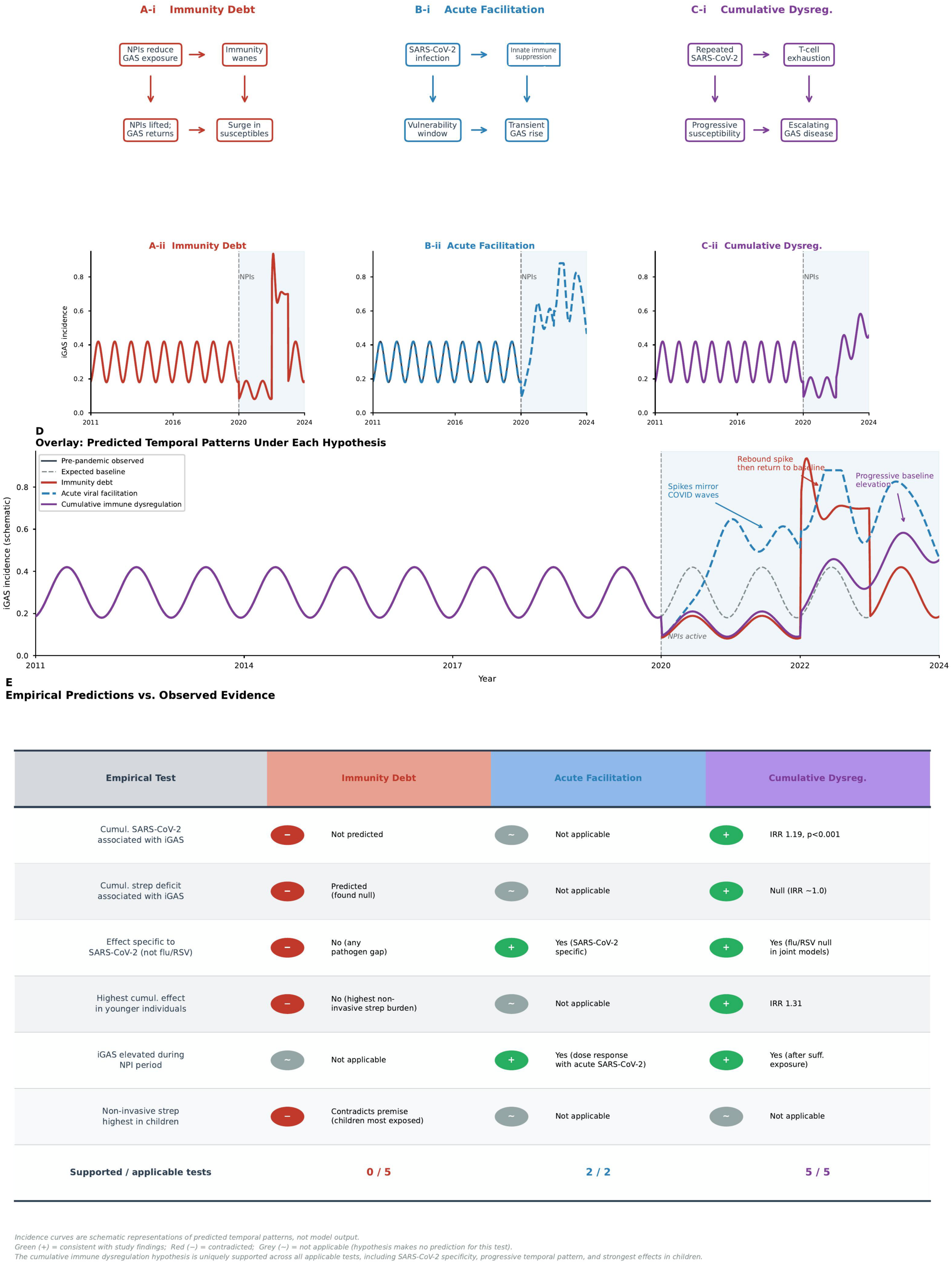
Competing Mechanistic Hypotheses for the Post-Pandemic Invasive Group A Streptococcal Disease Surge and Their Empirical Predictions. Panels A-i, B-i, and C-i illustrate the causal pathways posited by the immunity debt, acute viral facilitation, and cumulative immune dysregulation hypotheses, respectively. Panels A-ii, B-ii, and C-ii show schematic iGAS incidence trajectories predicted by each hypothesis relative to the pre-pandemic observed pattern (black) and expected secular baseline (dashed grey). Panel D overlays all three predicted trajectories on a single timeline. Panel E summarizes the consistency of each hypothesis with five pre-specified empirical tests. Green circles indicate findings consistent with the hypothesis; red circles indicate findings that contradict it; grey circles indicate tests not directly addressable by the hypothesis. The cumulative immune dysregulation hypothesis was supported by 5 of 5 applicable tests, including the specificity of effects to SARS-CoV-2 (with null associations for influenza and RSV in joint models), the dose-response relationship with cumulative burden, and the strongest effects in younger individuals. Incidence curves in panels A-ii through D are schematic representations of predicted temporal patterns and do not reflect model output. IRR, incidence rate ratio.

## Supplementary Appendix 3: Respiratory Virus Co-Circulation

Respiratory virus activity for non-SARS-CoV-2 viruses was obtained from the Respiratory Virus Detection Surveillance System (RVDSS)^2^, which reports weekly laboratory detections across sentinel surveillance laboratories in Ontario. SARS-CoV-2 activity during pandemic period 1 was based on test-adjusted case estimates derived from the Ontario Case Management System and Ontario Laboratory Information System. Viral exposure measures were normalized by dividing by standard deviation (SD) units within each virus type to allow comparison of activity levels on a common scale. For SARS-CoV-2, time-period-specific SD measures were used for pandemic period 1 and pandemic period 2.

**Figure S4** displays normalized weekly activity for RSV, influenza (combined), influenza A, influenza B, and (during the pandemic) SARS-CoV-2. Panel A shows the pre-pandemic period (2011-2020), during which all respiratory viruses displayed characteristic winter seasonal patterns with annual peaks. RSV and influenza co-circulated each winter, though their peak timing varied across seasons. Panel B shows the pandemic period (2020-2024). All non-SARS-CoV-2 respiratory viruses were profoundly suppressed during 2020-2021 due to non-pharmaceutical interventions, with gradual re-emergence beginning in late 2021. SARS-CoV-2 activity dominated the pandemic period, with multiple major waves. Influenza B activity remained minimal throughout the pandemic period; consequently, analyses during the pandemic period were conducted for combined influenza rather than by subtype, as effectively all influenza during this period was influenza A.

The co-circulation of SARS-CoV-2 with other respiratory viruses during pandemic period 2, particularly the moderate correlation between lagged influenza and SARS-CoV-2 exposure (Pearson ρ = 0.33), motivates the negative control analyses presented in **Supplementary Appendix 6.**

**Figure S4.**
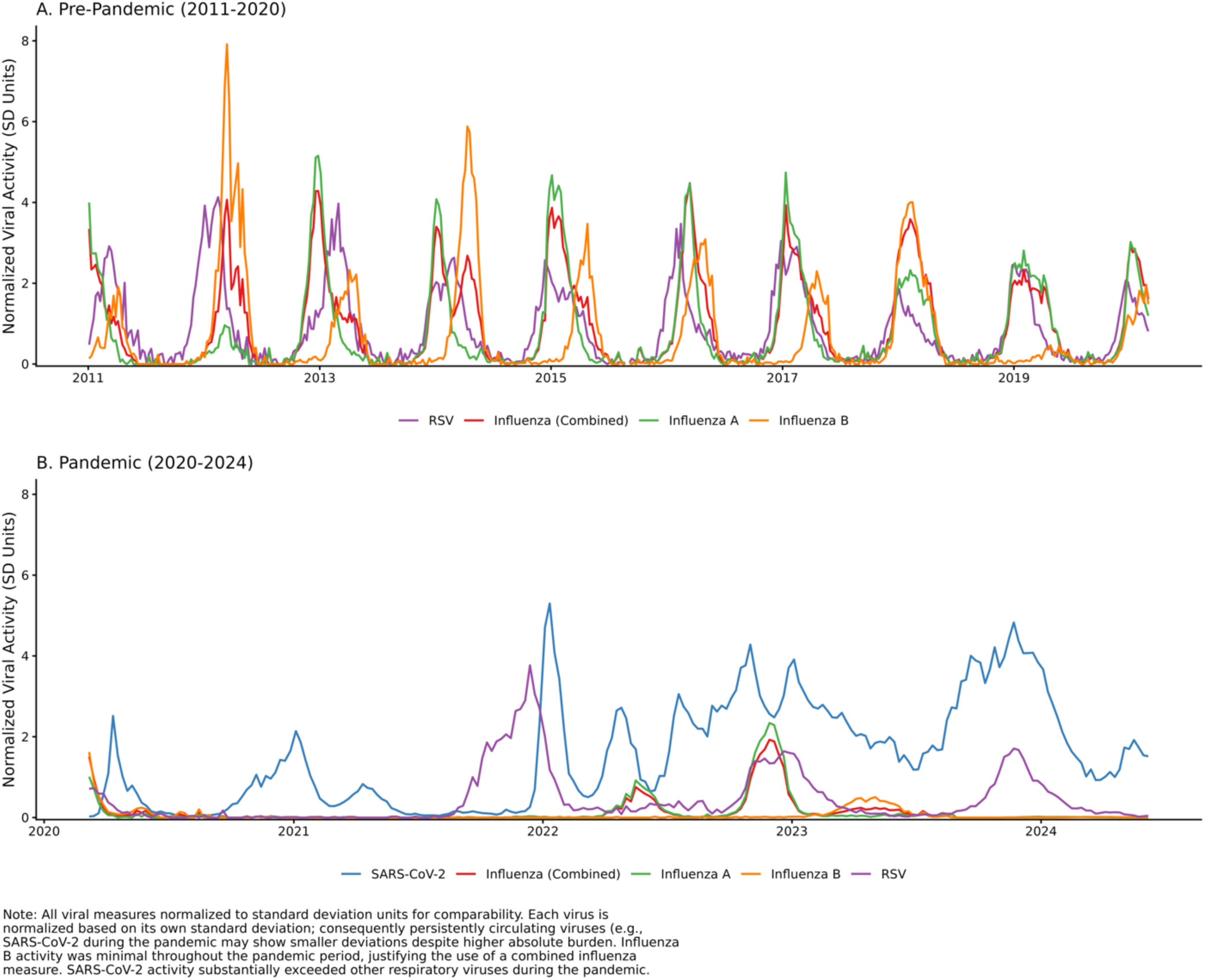
Respiratory virus co-circulation in the GTHA+ region, 2011-2024. (A) Pre-pandemic period showing seasonal patterns for RSV, influenza (combined, A, and B subtypes). (B) Pandemic period showing SARS-CoV-2, influenza, and RSV activity. All viral measures are normalized to standard deviation units for comparability; each virus is normalized to its own standard deviation, so persistently circulating viruses may show smaller deviations despite higher absolute burden.

## Supplementary Appendix 4: Cumulative Exposure Trajectories

To operationalize the cumulative SARS-CoV-2 burden hypothesis, we constructed running cumulative sums of weekly normalized viral exposures beginning at pandemic onset (March 2020). **Figure S5** displays these cumulative trajectories for SARS-CoV-2, influenza, RSV, and streptococcal disease.

Panel A shows cumulative viral exposures in normalized units. SARS-CoV-2 accumulated far more rapidly and to a substantially greater extent than influenza or RSV, reflecting both higher peak activity and more sustained circulation. By end of study, cumulative SARS-CoV-2 exposure exceeded 300 SD-units, compared with approximately 30 for influenza and 20 for RSV. This differential accumulation is central to the interpretation of cumulative burden models: the cumulative SARS-CoV-2 variable captures both the intensity and duration of population-level COVID-19 exposure.

Panel B shows cumulative streptococcal case counts (total strep, non-invasive, and iGAS). Non-invasive streptococcal disease accumulated steadily throughout the pandemic, with notable acceleration during pandemic period 2 coinciding with the iGAS surge, though iGAS represented a small fraction of total streptococcal disease throughout. The cumulative streptococcal trajectory is relevant to the immunity debt hypothesis test (**Supplementary** **Appendix 8**): if immunity debt were driving the iGAS surge, accumulated streptococcal deficit should predict declining iGAS risk.

**Figure S5.**
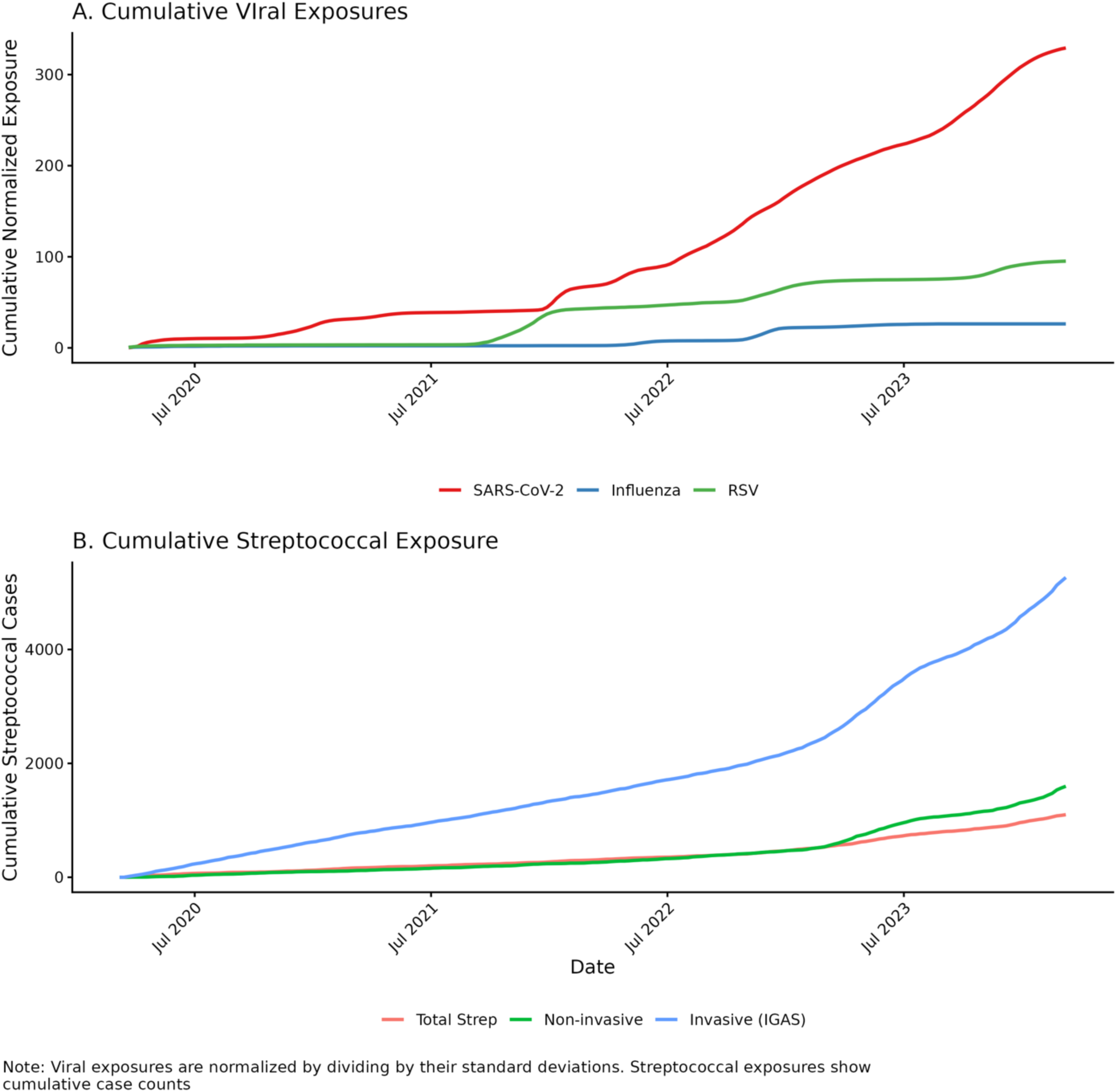
Cumulative exposure trajectories during the pandemic period (March 2020-June 2024). (A) Cumulative normalized viral exposures for SARS-CoV-2, influenza, and RSV. (B) Cumulative streptococcal case counts for total strep, non-invasive strep, and invasive GAS (iGAS).

## Supplementary Appendix 5: Age-Stratified SARS-CoV-2 Effects on Invasive Group A Streptococcal Disease

This appendix presents age-stratified associations between SARS-CoV-2 exposure and iGAS incidence. **Table S5a** shows acute effects from models including only 2-week lagged SARS-CoV-2 exposure. **Table S5b** shows cumulative effects from joint models including both acute and cumulative SARS-CoV-2 burden. Pandemic period 1 (March 2020-August 2022) was characterized by active non-pharmaceutical interventions; pandemic period 2 (September 2022-March 2024) followed relaxation of public health measures. All models were adjusted for sex, seasonality (Fourier transforms with annual periodicity), and quadratic time trend. Age-stratified models used standard negative binomial regression (nbreg command in Stata) with covariate adjustment for sex.

**Table S5a.**
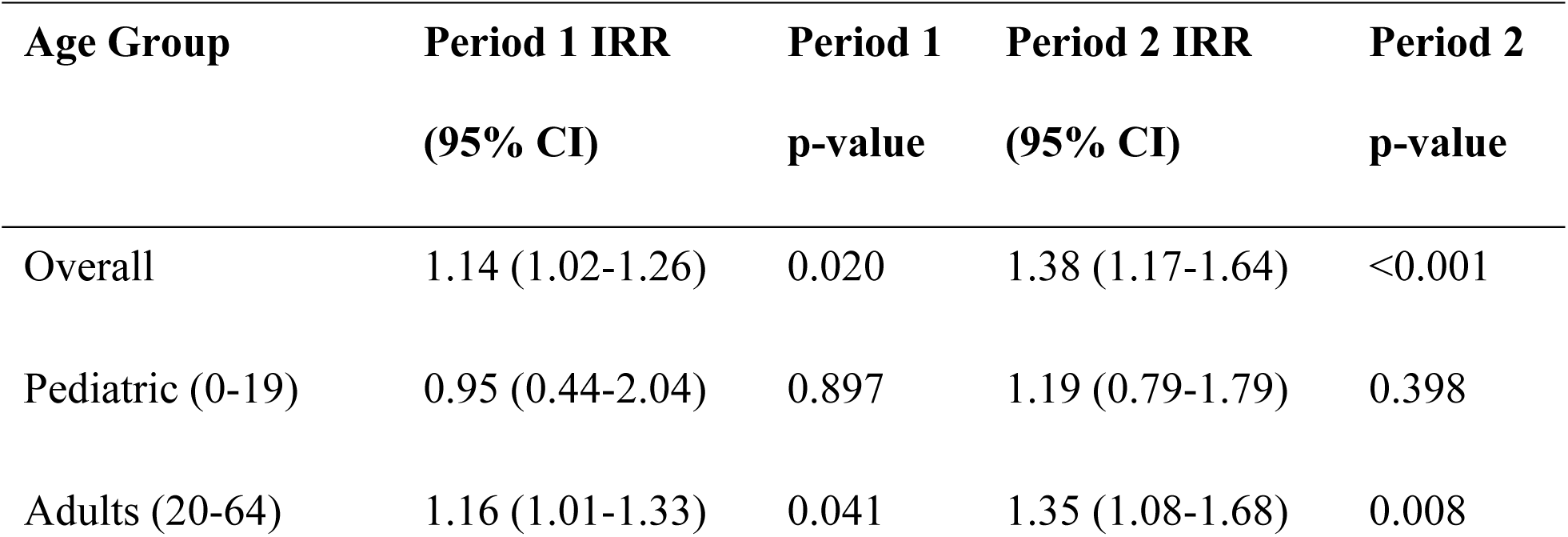
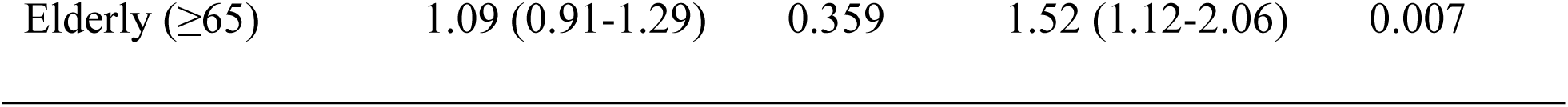
Acute SARS-CoV-2 effects on iGAS incidence, by age group and pandemic period (2-week lag, per 1-SD increase).

**Table S5b.**
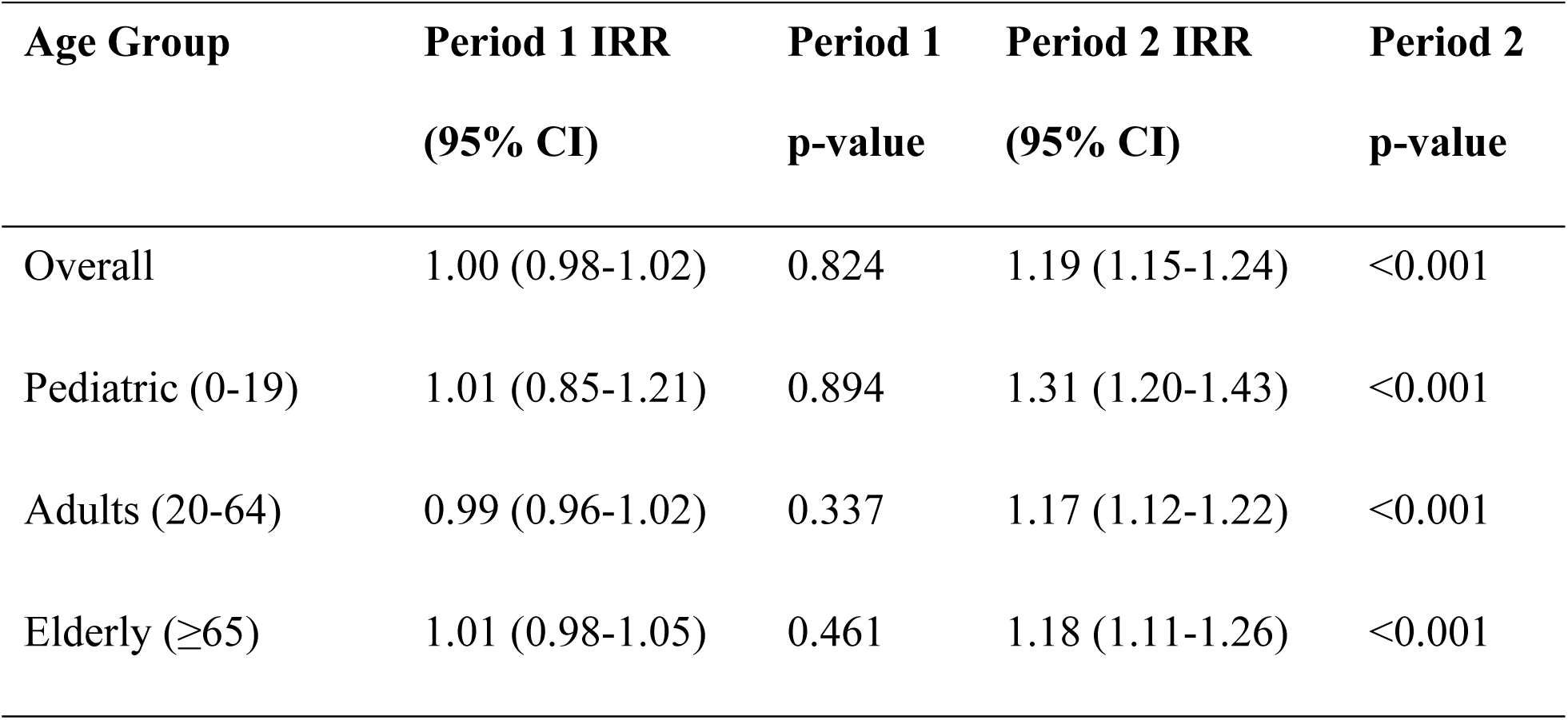
Cumulative SARS-CoV-2 effects on iGAS incidence, by age group and pandemic period (from joint acute + cumulative models).

IRR = incidence rate ratio; CI = confidence interval. Acute effects represent the association between 2-week lagged standardized SARS-CoV-2 exposure (per 1-SD increase) and iGAS incidence. Cumulative effects represent the association between cumulative standardized SARS-CoV-2 burden (running sum from March 2020) and iGAS incidence, adjusted for acute effects. During pandemic period 2, models including cumulative burden showed substantially better fit than acute-only models (overall population: ΔAIC = −157.5).

## Supplementary Appendix 6: Negative Control Analyses

To evaluate the specificity of observed SARS-CoV-2 associations with iGAS, we applied identical model structures to influenza (combined, A, and B subtypes) and respiratory syncytial virus (RSV). All models used 2-week lagged, population-normalized viral exposure measures (per 1-SD increase) and were adjusted for age group, sex, and quadratic time trends, with population offset. Seasonal adjustment used Fourier harmonic terms (sine and cosine).

### Pre-pandemic period (March 2011-February 2020)

In the pre-pandemic period, all respiratory viruses showed significant apparent associations with iGAS in models without seasonal adjustment (**Table S6.1**). After inclusion of Fourier harmonic terms, all associations became null, indicating that previously observed virus-iGAS associations were attributable to shared winter seasonality rather than direct viral effects.

**Table S6.1.** Association between respiratory virus circulation and iGAS incidence, pre-pandemic period, with and without seasonal adjustment.

| **Virus** | **Seasonal**<br>**Harmonics** | **IRR (95% CI)** | **p-**<br>**value** | **AIC** |
| --- | --- | --- | --- | --- |
| RSV | No | 1.21 (1.14-1.27) | <0.001 | 5705.2 |
| RSV | Yes | 1.00 (0.91-1.10) | 0.950 | 5666.6 |
| Influenza | No | 1.16 (1.11-1.21) | <0.001 | 5706.3 |
| Influenza | Yes | 0.96 (0.90-1.04) | 0.313 | 5665.6 |
| Influenza A | No | 1.11 (1.07-1.16) | <0.001 | 5727.7 |
| Influenza A | Yes | 0.96 (0.90-1.02) | 0.136 | 5664.3 |
| Influenza B | No | 1.12 (1.08-1.17) | <0.001 | 5720.0 |
| Influenza B | Yes | 1.02 (0.97-1.07) | 0.448 | 5666.0 |

### Pandemic Period 1 (March 2020-August 2022)

During pandemic period 1, neither acute nor cumulative influenza or RSV exposure was significantly associated with iGAS incidence (Table S6.2).

**Table S6.2.** Association between respiratory virus circulation and iGAS incidence, pandemic period 1.

| **Virus** | **Model** | **Acute IRR**<br>**(95% CI)** | **p-**<br>**value** | **Cumulative**<br>**IRR (95% CI)** | **p-**<br>**value** | **AIC** |
| --- | --- | --- | --- | --- | --- | --- |
| Influenza | Acute | 1.12 (0.74-1.69) | 0.593 | — | — | 1188.6 |
| Influenza | Joint | 0.90 (0.35-2.28) | 0.821 | 1.05 (0.93-1.18) | 0.413 | 1182.2 |
| RSV | Acute | 1.15 (0.99-1.35) | 0.072 | — | — | 1185.8 |
| RSV | Joint | 1.13 (0.96-1.33) | 0.142 | 1.01 (0.99-1.02) | 0.374 | 1179.2 |

### Pandemic Period 2: Acute Effects

Acute RSV exposure showed no association with iGAS in Period 2 (**Table S6.3**). Acute influenza showed a borderline association (IRR 1.31, 95% CI 0.99-1.73; p = 0.058) that became nominally significant when jointly modeled with acute SARS-CoV-2 (IRR 1.42, 95% CI 1.05-1.92; p = 0.023), while SARS-CoV-2 became non-significant (IRR 1.28, 95% CI 0.91-1.81; p = 0.160). The Pearson correlation between lagged influenza and SARS-CoV-2 exposure in period 2 was 0.33 (Spearman ρ = 0.26, p < 0.001), consistent with co-circulation and cautioning against joint models of these two exposures in the acute framework.

**Table S6.3.** Acute respiratory virus associations with iGAS incidence, pandemic period 2.

| Virus | Adjustment | Virus IRR<br>(95% CI) | p-value | SARS-CoV-2<br>IRR (95% CI) | p-value | AIC |
| --- | --- | --- | --- | --- | --- | --- |
| Influenza | Unadjusted | 1.31 (0.99-1.73) | 0.058 | — | — | 918.3 |
| Influenza | SARS-CoV-2 adjusted | 1.42 (1.05-1.92) | 0.023 | 1.28 (0.91-1.81) | 0.160 | 918.3 |
| RSV | Unadjusted | 1.00 (0.72-1.37) | 0.977 | — | — | 1674.7 |

### Pandemic Period 2: Cumulative Effects

Cumulative influenza burden showed no association with iGAS when modeled independently (IRR 0.99, 95% CI 0.88-1.11; p = 0.831) (Table S6.4). Cumulative RSV was significantly associated with iGAS when modeled alone (IRR 1.31, 95% CI 1.24-1.39; p < 0.001); however, this association was eliminated in head-to-head models with cumulative SARS-CoV-2 (Table S6.5).

**Table S6.4.** Cumulative respiratory virus associations with iGAS incidence, pandemic period 2.

| **Virus** | **Acute IRR**<br>**(95% CI)** | **p-**<br>**value** | **Cumulative**<br>**IRR (95% CI)** | **p-**<br>**value** | **AIC** |
| --- | --- | --- | --- | --- | --- |
| Influenza | 1.34 (0.93-1.93) | 0.114 | 0.99 (0.88-1.11) | 0.831 | 920.2 |
| RSV | 0.50 (0.36-0.71) | <0.001 | 1.31 (1.24-1.39) | <0.001 | 1568.6 |
| SARS-CoV-2 | — | — | 1.19 (1.15-1.24) | <0.001 | 1505.4 |

### Specificity Analyses: Competing Exposures and Secular Trends

In models jointly including cumulative SARS-CoV-2 and cumulative RSV, the SARS-CoV-2 association persisted (IRR 1.19, 95% CI 1.14-1.25; p < 0.001) while RSV became null (IRR 1.02, 95% CI 0.94-1.11; p = 0.626) (Table S6.5). Cumulative SARS-CoV-2 was also robust to inclusion of a period 2-specific linear time trend (IRR 1.18, 95% CI 1.13-1.23; p < 0.001); the time trend itself was non-significant (IRR 1.14, 95% CI 0.97-1.32; p = 0.108), indicating that the cumulative SARS-CoV-2 association is not explained by a general secular increase in iGAS risk during this period.

**Table S6.5.**
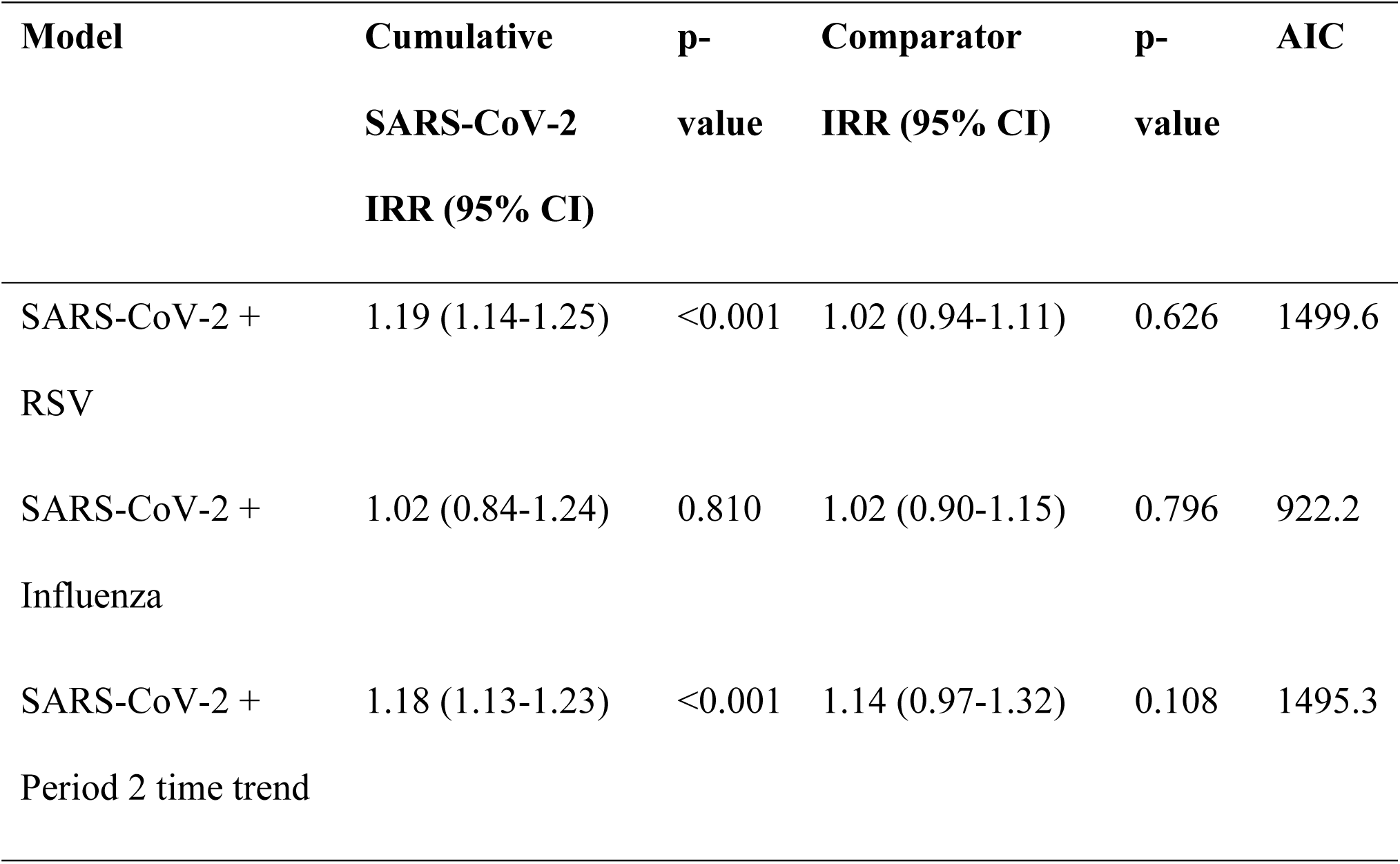
Head-to-head models: cumulative SARS-CoV-2 versus cumulative respiratory viruses and linear time trend, pandemic period 2.

## Supplementary Appendix 7: Non-Invasive Streptococcal Disease and Propensity to Invade

To characterize whether SARS-CoV-2 exposure was associated with increased streptococcal disease broadly or specifically with invasive disease, we conducted two complementary analyses. First, we modeled non-invasive streptococcal infections (pharyngitis, cellulitis, erysipelas, and other non-invasive presentations) as the outcome with population as offset, using the same model structure as the primary iGAS analyses. Second, we modeled iGAS incidence with total streptococcal disease (invasive + non-invasive + 0.5) as offset rather than population, thereby estimating the propensity for streptococcal infections to become invasive conditional on SARS-CoV-2 exposure.

### Non-Invasive Streptococcal Infections

During pandemic period 1, neither acute nor cumulative SARS-CoV-2 exposure was associated with non-invasive streptococcal incidence (**Table S7.1**). During pandemic period 2, acute SARS-CoV-2 was positively associated with non-invasive strep (IRR 1.25, 95% CI 1.06-1.48; p = 0.010), and cumulative SARS-CoV-2 showed a strong association (IRR 1.22, 95% CI 1.18-1.25; p < 0.001) of nearly identical magnitude to its effect on iGAS (IRR 1.19, 95% CI 1.15-1.24; p < 0.001) (**Table S7.2**).

**Table S7.1.**
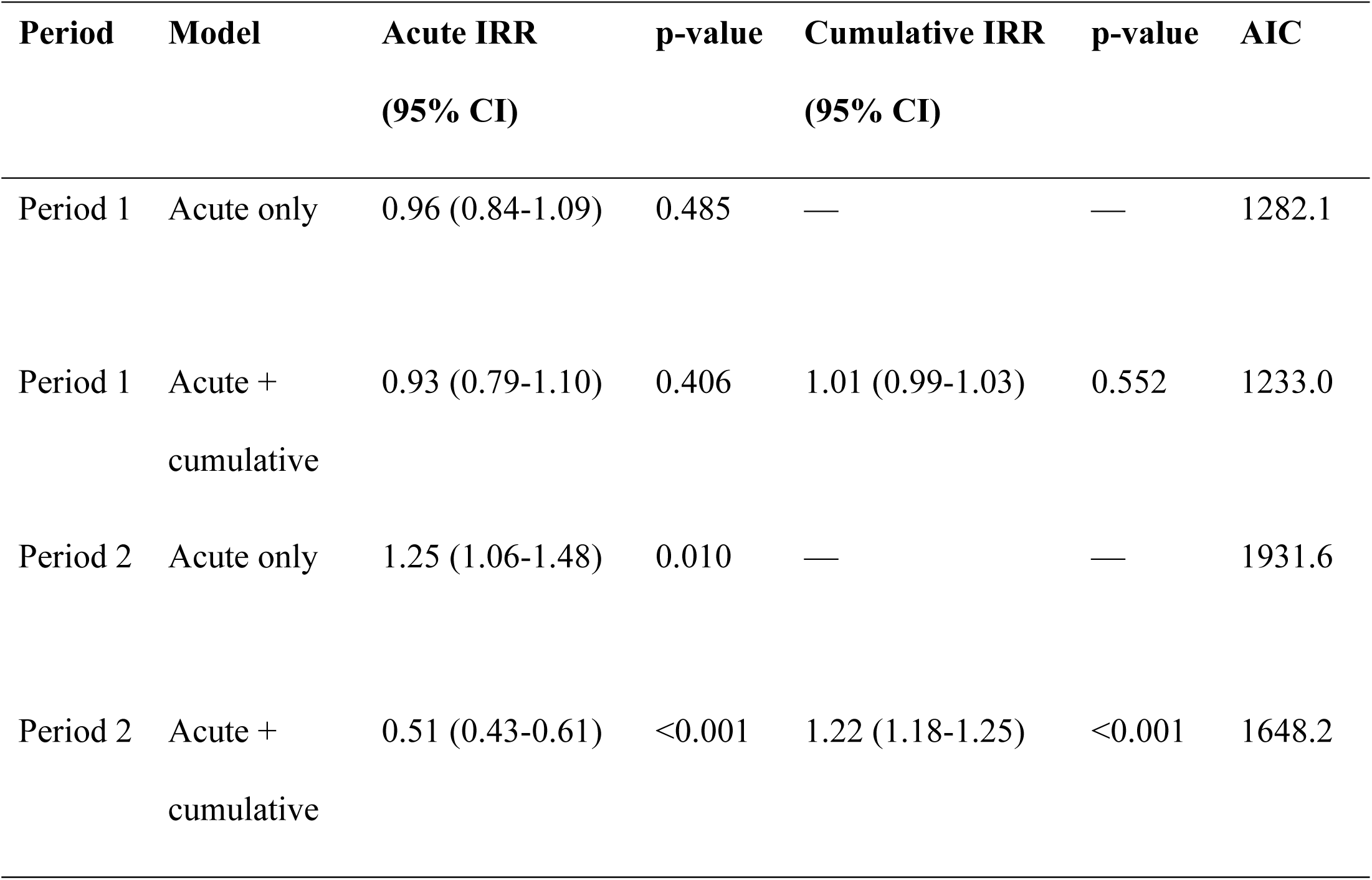
Association between SARS-CoV-2 exposure and non-invasive streptococcal disease incidence, by period and model specification.

**Table S7.2.**
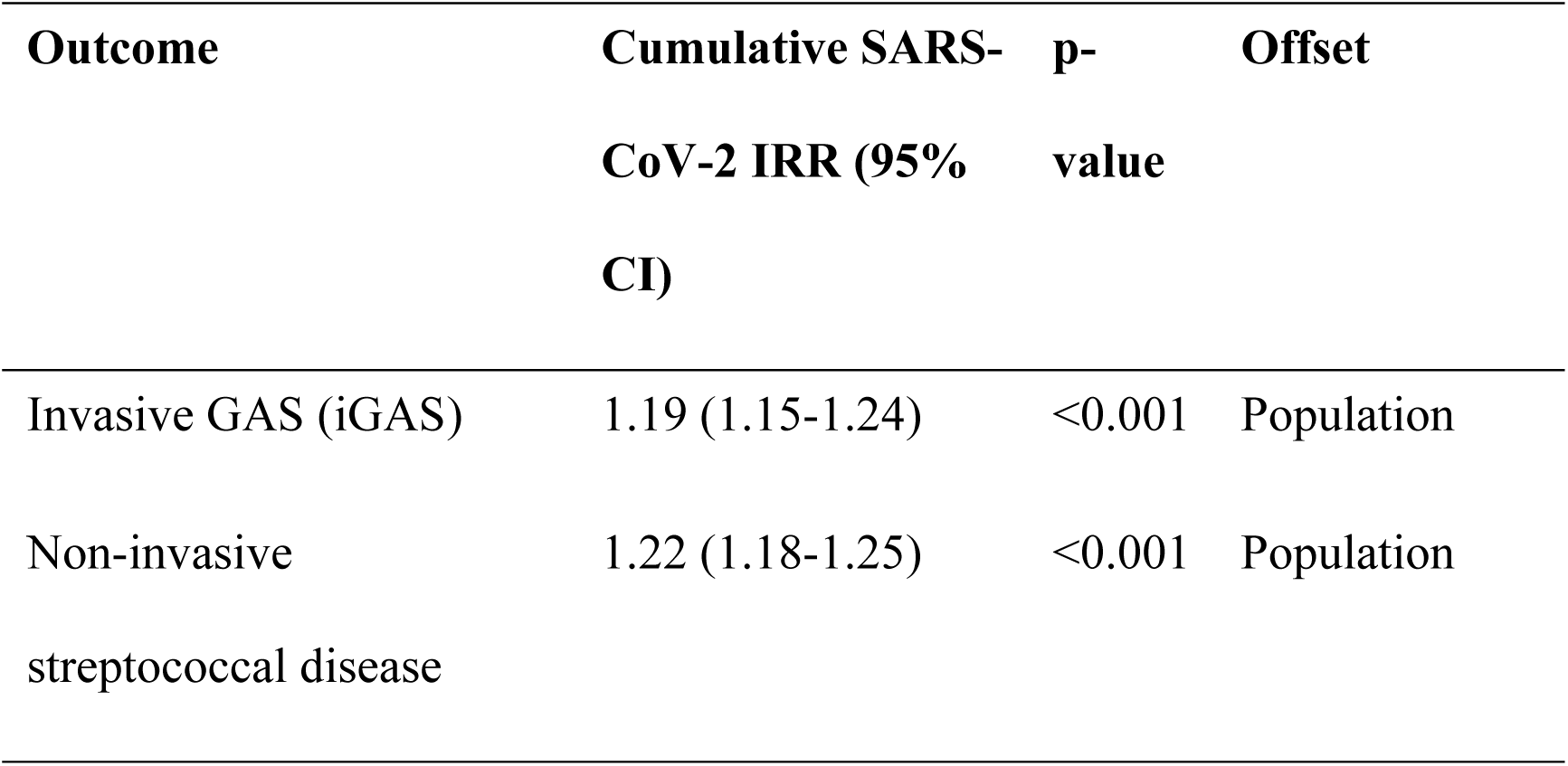
Comparison of cumulative SARS-CoV-2 effects on invasive and non-invasive streptococcal disease, pandemic period 2.

### Propensity to Invade

When total streptococcal disease was used as the offset (modeling iGAS per streptococcal infection rather than per capita), SARS-CoV-2 showed a borderline acute association during pandemic period 1 (IRR 1.12, 95% CI 1.00-1.25; p = 0.047 in the acute-only model; IRR 1.14, 95% CI 1.01-1.29; p = 0.036 in the joint model) but no cumulative effect (**Table S7.3**). During pandemic period 2, neither acute nor cumulative SARS-CoV-2 exposure was associated with invasion propensity. These findings indicate that SARS-CoV-2 increases susceptibility to streptococcal disease broadly rather than specifically increasing the propensity for streptococcal infections to become invasive.

**Table S7.3.**
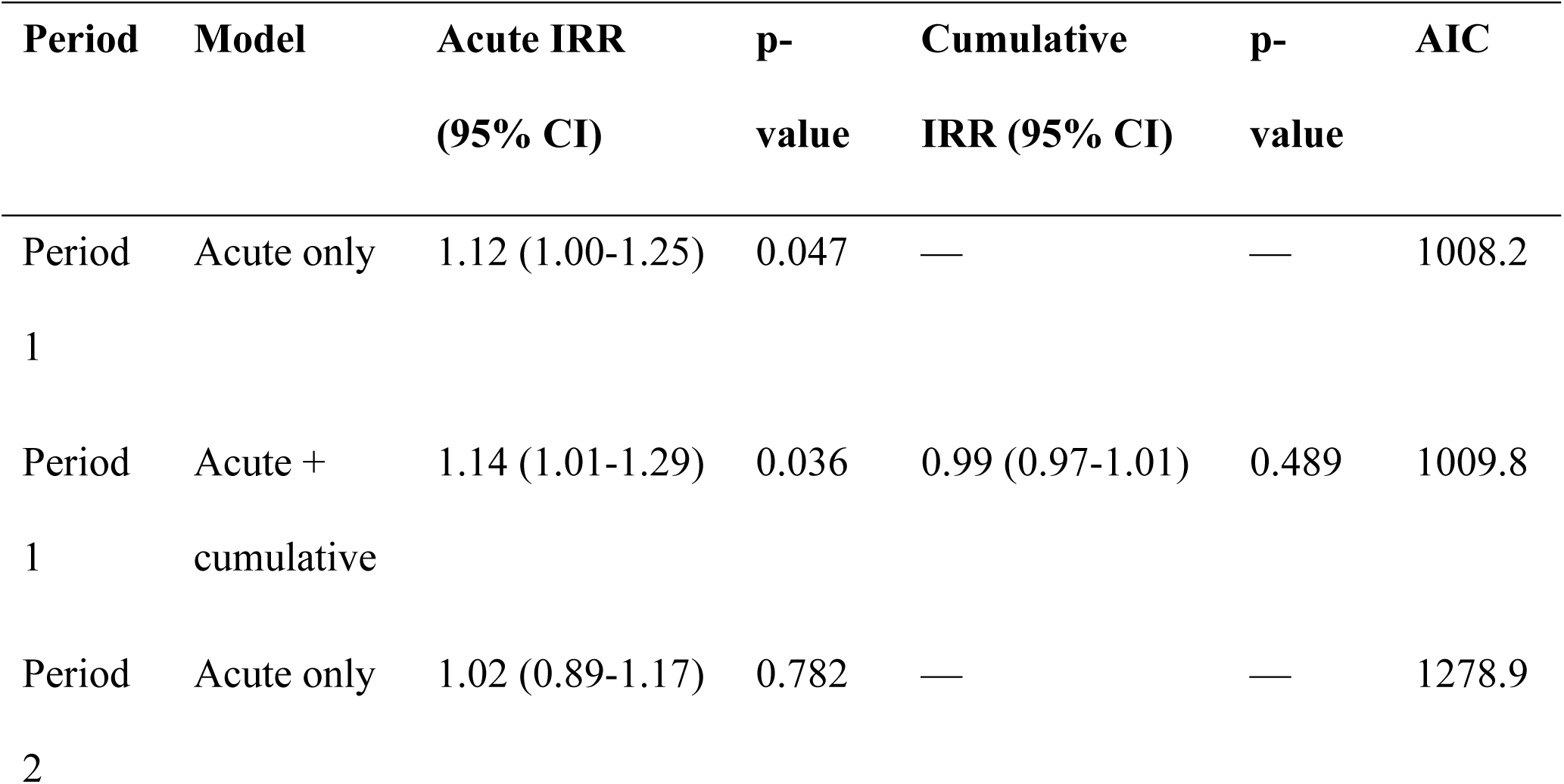
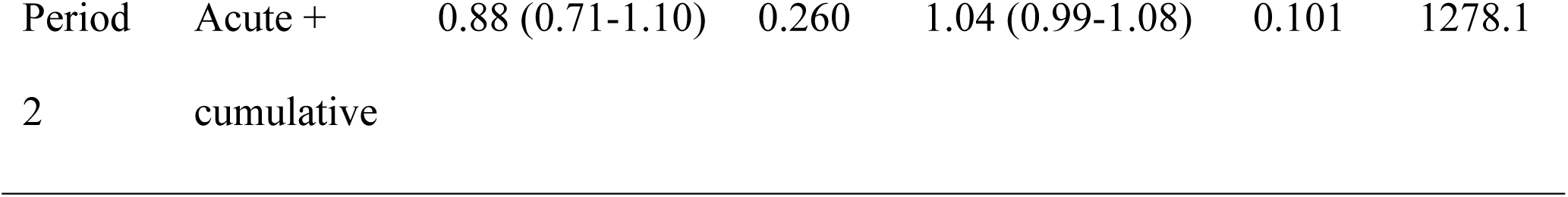
Association between SARS-CoV-2 exposure and propensity for streptococcal invasion (iGAS incidence with total streptococcal disease as offset).

## Supplementary Appendix 8: Immunity Debt Hypothesis Testing

The immunity debt hypothesis posits that reduced pathogen circulation during the pandemic resulted in a population-level deficit in immune boosting, increasing susceptibility to infection upon pathogen re-emergence. Under this hypothesis, accumulated streptococcal deficit,rather than SARS-CoV-2 exposure,would be the primary driver of the post-pandemic iGAS surge. We tested this by including cumulative all-strep burden (invasive + non-invasive) alongside cumulative SARS-CoV-2 in pandemic period 2 models. If immunity debt were the dominant mechanism, cumulative streptococcal burden should show a negative association with iGAS (less prior exposure → greater current susceptibility) and should attenuate the cumulative SARS-CoV-2 effect.

Cumulative SARS-CoV-2 was strongly and consistently associated with iGAS across all age groups, with no attenuation upon inclusion of cumulative streptococcal burden (Table S8.1). Cumulative streptococcal burden showed no association with iGAS overall (IRR 1.00, 95% CI 1.00-1.00; p = 0.730) or among children (IRR 1.00, 95% CI 0.99-1.01; p = 0.620). Among adults (IRR 1.01, 95% CI 1.00-1.02; p = 0.011) and older adults (IRR 1.01, 95% CI 1.00-1.03; p = 0.055), cumulative streptococcal burden showed a modest positive association,the opposite direction predicted by immunity debt, in which reduced streptococcal exposure should increase susceptibility. These positive associations are consistent with greater streptococcal circulation providing more opportunities for invasion rather than immunity debt-mediated susceptibility.

**Table S8.1.** Immunity debt hypothesis test: cumulative SARS-CoV-2 and cumulative streptococcal burden as joint predictors of iGAS incidence, pandemic period 2. All models adjusted for acute SARS-CoV-2 exposure, seasonal harmonics, quadratic time trend, and sex (age-stratified models) or age and sex (overall model), with population offset.

| **Age Group** | **Cumulative SARS-CoV-2 IRR (95% CI)** | **p-value** | **Cumulative All-Strep IRR (95% CI)** | **p-value** | **AIC** |
| --- | --- | --- | --- | --- | --- |
| Overall | 1.19 (1.15-1.24) | <0.001 | 1.00 (1.00-1.00) | 0.730 | 1507.3 |
| Pediatric (0-19) | 1.31 (1.20-1.43) | <0.001 | 1.00 (0.99-1.01) | 0.620 | 372.4 |
| Adults (20-64) | 1.14 (1.09-1.20) | <0.001 | 1.01 (1.00-1.02) | 0.011 | 585.8 |
| Elderly ($\geq 65$) | 1.17 (1.09-1.25) | <0.001 | 1.01 (1.00-1.03) | 0.055 | 524.8 |

